# Initial findings from the DecodeME genome-wide association study of myalgic encephalomyelitis/chronic fatigue syndrome

**DOI:** 10.1101/2025.08.06.25333109

**Authors:** Genetics Delivery Team, Thibaud Boutin, Andrew D. Bretherick, Joshua J. Dibble, Esther Ewaoluwagbemiga, Emma Northwood, Gemma L. Samms, Veronique Vitart, Project and Cohort Delivery Team, Øyvind Almelid, Tom Baker, Malgorzata Clyde, Anne Connolly, Diana Garcia, Shona M. Kerr, Claire Tripp, Jareth C. Wolfe, Patient and Public Involvement, Jackie Goold, Gemma Hoyes, Sian Leary, Simon J. McGrath, Julie Milton, Anna Redshaw, Jim M. Wilson, Marketing and Communications Team, Helen Baxter, Danielle Boobyer, Claire Dransfield, Daphne Lamirel, Isabel Lewis, Nina Muirhead, Ella Ponting, Charles Shepherd, Alice Turner, University of Edinburgh Team, Sumy V. Baby, Sjoerd Beentjes, John Ireland, Ava Khamseh, Ewan McDowall, David Perry, Joshua Slaughter, Genetic Epidemiology of ME/CFS Consortium, Erik Abner, Cindy G. Boer, Estonian Biobank Research Team, Sarah Finer, Genes & Health Research Team, Hele Haapaniemi, Hanna M. Ollila, Beth Pollack, Judith Rosmalen, Erika Romppanen, Sirine Saafi, Richa Saxena, Nasa Sinnott-Armstrong, Anniina Tervi, Lea Urpa, Jesse Valliere, David A. van Heel, Management Team, Sonya Chowdhury, Andy Devereux-Cooke, Chris P. Ponting

## Abstract

Myalgic encephalomyelitis / chronic fatigue syndrome (ME/CFS) is a common, poorly understood disease that has no effective treatments, and has long been underserved by scientific research and national health systems. It is a sex-biased disease towards females that is often triggered by an infection, and its hallmark symptom is post-exertional malaise. People with ME/CFS often report their symptoms being disbelieved. The biological mechanisms causing ME/CFS remain unclear. We recruited 21,620 ME/CFS cases and performed genome-wide association studies (GWAS) for up to 15,579 cases and 259,909 population controls with European genetic ancestry. In these GWAS, we discovered eight loci that are significantly associated with ME/CFS, including three near *BTN2A2, OLFM4*, and *RABGAP1L* genes that act in the response to viral or bacterial infection. Four of the eight loci (*RABGAP1L, FBXL4, OLFM4, CA10*) were associated at *p* < 0.05 with cases ascertained using post-exertional malaise and fatigue in the UK Biobank and the Netherlands biobank Lifelines. We found no evidence of sex-bias among discovered associations, and replicated in males two genetic signals (*ARFGEF2, CA10*) discovered in females. The ME/CFS association near *CA10* colocalises with a known association to multisite chronic pain. We found no evidence that the eight ME/CFS genetic signals share common causal genetic variants with depression or anxiety. Our findings suggest that both immunological and neurological processes are involved in the genetic risk of ME/CFS.

**LAY SUMMARY:** Myalgic encephalomyelitis / chronic fatigue syndrome (ME/CFS) is a common, disabling illness. It affects more females than males, and in most cases, starts after an infection. Little is known about the biological mechanisms that cause ME/CFS, despite many attempts to uncover them, and it has no effective treatments. To understand ME/CFS better, our study, DecodeME, compared the DNA of 15,579 people with ME/CFS with the DNA of 259,909 people without ME/CFS, all of European descent. DNA is a molecule that makes up our genes. Our genes make many different molecules called proteins, each of which does very specific things in the body. Finding variations in genes that differ between people with or without a disease can therefore point to what causes it. We found that people with ME/CFS are more likely to carry certain DNA differences in eight regions of their genome, and so these variants tell us about possible biological causes of ME/CFS. However, as these differences are also often found in people without ME/CFS they cannot cleanly separate who is at risk and who is not, and therefore do not provide a definitive test. Most of these regions contain several genes. Our methods did not allow us to conclusively locate the ones most relevant to ME/CFS in each region, but public data allowed us to pick out the most likely ones. Three of the most likely genes produce proteins that respond to an infection. Another likely gene is related to chronic pain. None are related to depression or anxiety. We found nothing to explain why more females than males get ME/CFS. Overall, DecodeME shows that ME/CFS is partly caused by genes related to the immune and nervous systems.

## INTRODUCTION

An estimated 67 million people worldwide live with myalgic encephalomyelitis / chronic fatigue syndrome (ME/CFS) symptoms (1). Approximately 80% of them are female (2,3) and, in one survey, about half were housebound or bedbound (4). A key symptom of ME/CFS is post-exertional malaise. This is a worsening of symptoms, both physical and cognitive, after even minor physical or mental exertion that an individual would have tolerated easily when well (5). The onset of post-exertional malaise is typically delayed by 12 to 48 hours after the triggering activity, and can last for days or even weeks, sometimes leading to a relapse. Other symptoms include pain and profound fatigue unrelieved by sleep (2).

About 10% of people with infectious diseases such as glandular fever (infectious mononucleosis) have ME/CFS symptoms within 12 months (6,7), and 13%–45% of people with Long Covid meet ME/CFS diagnostic criteria (8). There is no positive diagnostic test for ME/CFS, no known cause, and while it can relapse and remit, full recovery is rare, at about 5% (9,10).

Identifying genetic risk factors for ME/CFS is a high research priority for people with the illness and their carers (11) and is feasible because predisposition to ME/CFS is partly heritable (12,13). ME/CFS is not a monogenic disease (14), but rather a multifactorial one whose genetics is best studied using a genome-wide association study (GWAS) design.

Unlike other complex diseases, ME/CFS research has not yet benefited from GWAS. This is likely because previously studied cohorts have not been large enough (15) to identify multiple disease-associated loci. Studies with fewer than 1,000 cases have failed to identify susceptibility loci with any confidence (16,17). Studies with approximately 2,000 ME/CFS cases from the UK Biobank (UKB) have also not yielded robust associations (18–21). This could partly reflect the UK Biobank’s bias towards more healthy volunteers (22). It could also be due to these studies’ reliance on self-reported diagnoses or on a hospital episode statistics (ICD10) diagnosis code that is neither specific to ME/CFS nor accurate (23).

Here, we present the genetic results of DecodeME, a large ME/CFS GWAS co-produced by people with ME/CFS and their carers, and scientists (24). We aimed to identify common genetic variants associated with ME/CFS by comparing DNA that we collected from UK residents with clearly defined ME/CFS, with that of population controls from the UKB (25). Our goal was to find genetic risk factors that point to biomolecular mechanisms of ME/CFS and that could ultimately lead to diagnostics and effective treatment of this debilitating condition.

Given ME/CFS’s female predominance and frequent infectious trigger, we also analysed females (those assigned female at birth) and males (those assigned male at birth) together and then separately and stratified by whether disease onset was infectious or not. These GWAS yielded eight genome-wide significant associations, which indicate a genetic contribution, and an immunological and neurological basis, to this poorly understood disease.

## METHODS

### Participant recruitment and data collection

DecodeME was planned and delivered as a patient and public involvement (PPI) co-production with researchers (24). People with lived experience of ME/CFS were an integral part of the team that led the recruitment of participants via social and traditional media. We recruited participants between 12 September 2022 and 31 January 2024. We used podcasts, webinars, blog posts, media interviews, social media, and articles in ME/CFS charity members’ print magazines to build trust and understanding about DecodeME and to recruit to the study.

Participants completed a phenotype questionnaire which they could complete either online or on paper. We gave the most severely ill telephone support, if needed. We made it as easy as possible for people with ME/CFS, especially those housebound or bedbound, to give us their DNA, by using a ‘spit and post’ design to collect saliva samples (**Supplementary Methods**).

### Genotyping and analysis

Thermo Fisher genotyped DNA from cases using the UKB Axiom^TM^ array, the same platform used to genotype the UKB participants who were used as controls in this study. We published the project’s Data Analysis Plan online in March 2023 and updated it in March 2024, before we received case-genotype data. We first performed data and sample quality control (QC). We then merged case and control genotype data to perform a joint imputation, using a reference panel from the whole-genome sequences of more than 200,000 gender-matched UKB samples (**Supplementary Methods**).

We then compared case genotypes with those from UKB controls in a GWAS using a genome-wide significance threshold of *p* < 5 x 10^−8^ for variants with a minor allele frequency (MAF) ≥ 1% (**Supplementary Methods**). We imputed human leukocyte antigen (HLA) alleles separately using the HLA*IMP:02 algorithm, which the UKB also used for this study’s control individuals (25). Further methodological detail is provided in the **Supplementary Methods**.

### Case and control definitions

For genetic analyses, we used case inclusion and exclusion criteria based on the Canadian Consensus criteria and/or US Institute of Medicine / National Academy of Medicine criteria for ME/CFS (5,26). Both require post-exertional malaise as a symptom (**Supplementary Methods**). To meet these criteria, we did not include participants as cases if they reported that their illness had started within the previous six months, or had lasted their whole life. Additionally, cases needed to live in the UK, be at least 16 years old, and to have received a diagnosis of ME, CFS, ME/CFS or CFS/ME from a health professional. Also, those with an ME/CFS diagnosis after SARS-CoV-2 infection needed not to have been hospitalised, nor to have heart or lung damage, because of this infection.

We selected general population controls from UKB participants genotyped with the same Axiom^TM^ array if they had no associated evidence for ME/CFS, had no discrepancies between self-reported gender and genetically determined sex, did not present with a sex chromosome aneuploidy, were not heterozygosity outliers, and were in the list of UKB phased samples (**Supplementary Methods**). In analyses, controls were sampled to match the female-to-male ratio and genetic ancestry of DecodeME cases.

### Association testing

We performed genome-wide association testing using REGENIE (27), a machine-learning approach to whole-genome regression on phenotypes using data from a large number of individuals, for: (i) all DecodeME cases versus selected UKB controls; (ii) DecodeME cases versus UKB controls separately for females and males; and, (iii) DecodeME cases versus UKB controls separately for cases reporting an infection at onset and those not reporting an infection at onset. We defined infection-onset cases as those who answered ‘glandular fever’, ‘COVID-19’ or ‘another infection’ to the question, ‘Did you have an infection when, or just before, your first ME/CFS symptoms started?’ We defined non-infection-onset cases as those who answered ‘No’ or ‘Don’t know’ to this question.

We used the Firth logistic regression implemented in REGENIE, which is robust to case-control imbalance. The approach implemented in REGENIE also allowed us to account for covariates such as sex and genetic ancestry, using principal components (PCs) that explained most of the inter-individual variation not due to ME/CFS case status. Not accounting for ancestry could influence ME/CFS risk-association and/or confound case-control genetic associations. In the REGENIE modelling, we also fitted a polygenic random effect that accounted for cryptic and non-cryptic relatedness.

For REGENIE analyses, cases and controls were all of European genetic ancestry. We fitted sex (where appropriate) and the first 20 genetic PCs as covariates (28) (**Supplementary Methods**). We tested variants on chromosomes 1-22, reporting only those with imputation INFO scores ≥ 0.90 and MAF ≥ 1%.

Association case-control testing among Europeans of HLA alleles with AF > 1% used a logistic model, *glm()*, implemented in *R v4.4*, with genetic sex and the first 20 PCs as covariates (29).

### Post-GWAS analysis

We used FUMA (v1.8.0) to annotate genetic associations (30). FUMA is an integrative web platform that performs extensive functional annotation for DNA variants in genomic areas identified by lead variants using multiple resources.

To test for significant colocalisation between the FUMA-defined ME/CFS-associated genomic intervals and a second trait, we used the *R* package *coloc* v5.2.3 (31) to calculate the approximate Bayes factor posterior probability that both traits are associated and share a single causal variant (PP.H4.abf, or PPH4).

We investigated colocalisation between ME/CFS and gene expression by considering all protein-coding genes with one or more GTEx-v10 expression quantitative trait loci (eQTLs) present within the FUMA-defined ME/CFS-associated genomic intervals. We then tested for colocalisation with ME/CFS, as described above, for each of the 50 GTEx-v10 tissues under the single causal variant assumption (32).

We used LDSC (33) to estimate the genetic correlation between the phenotype of interest (ME/CFS) with other traits. We also used LDSC to estimate liability-scale SNP-based heritability (*h_l_*^2^).

To seek replication, we examined data obtained from nine large biobanks (**Supplementary Methods**). Specifically, we examined whether the 25 most significant variants found in DecodeME’s primary GWAS (at a less stringent threshold, *p* ≤ 9.4 x 10^−7^) were significantly associated with ME/CFS-related traits after accounting for multiple tests. We defined these traits as requiring cases to have reported post-exertional malaise and fatigue, in two cohorts – Lifelines, in the Netherlands (34)), and the UKB. In seven large cohorts in the USA, Estonia, Finland and the UK, we were only able to apply a wider case definition, including people with linked electronic health record data and/or self-report information consistent with ME/CFS and/or post-viral fatigue syndrome (**Supplementary Methods**).

## RESULTS

### The DecodeME cohort

Through its co-production model, DecodeME earned the trust and participation of people with ME/CFS in the UK and benefited from learning about their experiences of living with this disease. After launch, 26,901 people (84% female) completed the online or paper participant-entry questionnaire and consented to take part. Of these, 21,620 (85% female) met our study criteria of having a diagnosis of ME/CFS from a health professional, having post-exertional malaise as a symptom, and having symptoms consistent with Canadian Consensus and/or Institute of Medicine / National Academy of Medicine diagnostic criteria (5,26) (**Supplementary Methods**). We sent all cases a saliva DNA collection kit (**Fig. 1A**).

**Fig. 1:**
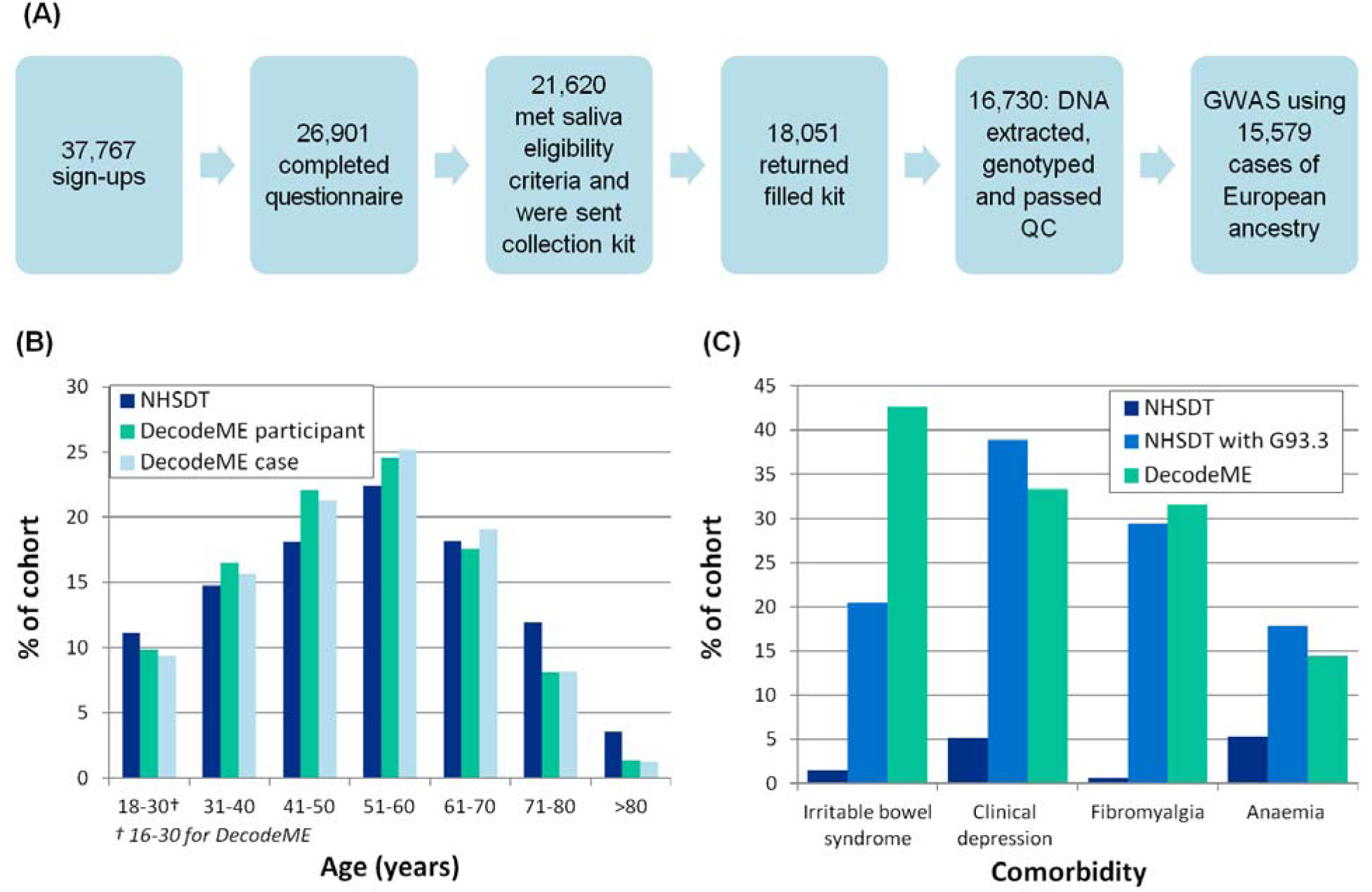
DecodeME cohort and characteristics. (A) DecodeME DNA cohort recruitment and genotyping process. (B) Age comparison of DecodeME participants (those completing the participant-entry questionnaire) with DecodeME GWAS-1 cases and with NHS England patients who had electronic healthcare records linked to the ICD-10 code G93.3 (“Postviral fatigue syndrome”), acquired from NHS DigiTrials (NHSDT). (C) Percentages of NHSDT patients with each of four common ME/CFS comorbidities, compared to both NHSDT patients with a linked G93.3 code, and DecodeME GWAS-1 cases.

We received samples from 18,051 DecodeME cases (84% female). Following genotyping on UK Biobank Axiom^TM^ arrays and quality-control steps (**Supplementary Methods**), Thermo Fisher generated genotype data for 18,266 samples, including duplicates.

To assess our cohort’s representativeness, we compared our DecodeME participants to English patients with electronic healthcare record links to G93.3. This is the ICD-10 code (‘Postviral fatigue syndrome’) that best reflects ME/CFS symptoms (35). The DecodeME cohort was broadly similar in age to these patients but contained a smaller proportion of older people with ME/CFS (**Fig. 1B**), possibly because our recruitment model was mostly internet-based.

DecodeME’s female bias (84.5% female) is greater than for the English G93.3 cohort (79.4%). This may be due to females using social media more (36) and younger people with ME/CFS being more often female (3). Most cases (59.5%) had lived with ME/CFS symptoms for at least ten years (**Table 1**). The DecodeME cohort was disproportionately White (96.5%), possibly due to low rates of ME/CFS diagnosis for other ethnicities (3,37) and barriers to research participation (38) that the project did not overcome.

**Table 1.** DecodeME cohort characteristics.

| **DecodeME cases** | **Female** | **Male** | **Total** | **% of total** |
| --- | --- | --- | --- | --- |
| Genotyped and passed quality control | 14,140 | 2,590 | 16,730 | 100.0 |
| Median age (years) | 52 | 55 |  |  |
| Age quartiles 1 and 3 (years) | 41 / 62 | 43 / 65 |  |  |
| **Comorbidities** |  |  |  |  |
| Irritable bowel syndrome | 6,232 | 901 | 7,133 | 42.6 |
| Clinical depression | 4,883 | 696 | 5,579 | 33.3 |
| Fibromyalgia | 4,797 | 477 | 5,274 | 31.5 |
| Anaemia needing treatment or blood transfusions | 2,344 | 75 | 2,419 | 14.5 |
| **Severity of ME/CFS (National Institute for Health and Care Excellence definitions)** |  |  |  |  |
| Mild | 4,067 | 728 | 4,795 | 28.7 |
| Moderate | 8,187 | 1,544 | 9,731 | 58.2 |
| Severe | 1,776 | 297 | 2,073 | 12.4 |
| Very severe | 110 | 21 | 131 | 0.8 |
| **Self-reported ethnicity** |  |  |  |  |
| White | 13,647 | 2,499 | 16,146 | 96.5 |
| Other ethnicities | 493 | 91 | 584 | 3.5 |
| **Years of having ME/CFS** |  |  |  |  |
| 0.5–1 | 220 | 34 | 254 | 1.5 |
| 1–3 | 1,268 | 224 | 1,492 | 8.9 |
| 3–5 | 1,469 | 214 | 1,683 | 10.1 |
| 5–10 | 2,848 | 507 | 3,355 | 20.1 |
| > 10 | 8,335 | 1,611 | 9,946 | 59.5 |
| **Pattern of symptoms over time** |  |  |  |  |
| Getting worse | 2,397 | 475 | 2,872 | 17.2 |
| Relapsing and remitting | 1,542 | 286 | 1,828 | 10.9 |
| Fluctuating | 8,721 | 1,443 | 10,164 | 60.8 |
| Not much change | 1,106 | 310 | 1,416 | 8.5 |
| Getting better | 369 | 73 | 442 | 2.6 |
| **Symptoms** |  |  |  |  |
| Unrefreshing sleep | 13,739 | 2,523 | 16,262 | 97.2 |
| Activities are a little affected by fatigue | 748 | 131 | 879 | 5.3 |
| Activities are significantly affected by fatigue | 13,384 | 2,456 | 15,840 | 94.7 |
| Fatigue (lack of energy) is disabling | 13,168 | 2,337 | 15,505 | 92.7 |
| Muscle pain | 12,342 | 2,058 | 14,400 | 86.1 |
| Confusion or 'brain fog' | 13,401 | 2,402 | 15,803 | 94.5 |
| Symptoms get worse with stress | 12,151 | 2,056 | 14,207 | 84.9 |
| **Infection when, or just before, first ME/CFS symptoms started** |  |  |  |  |
| Glandular fever (infectious mononucleosis) | 2,311 | 410 | 2,721 | 16.3% |
| COVID-19 virus | 462 | 84 | 546 | 3.3% |
| Another infection | 6,053 | 1,160 | 7,213 | 43.1% |
| No infection | 2,259 | 370 | 2,629 | 15.7% |
| Don't know | 3,055 | 566 | 3,621 | 21.6% |

DecodeME participants reported being diagnosed with four common comorbidities in similar proportions to English G93.3 cases (3) (**Fig. 1C**), except that they were diagnosed more than twice as often with irritable bowel syndrome.

Most ME/CFS cases reported that the impact that their symptoms had on their daily lives in terms of work, education, mobility and activities of daily living placed them in the categories that the National Institute for Health and Care Excellence class as showing ‘mild’ or ‘moderate’ impact of symptoms on everyday functioning (39) (**Table 1**). However, these labels do not fully match their lived experience, because most genotyped cases described their fatigue as being disabling (92.7%), had muscle pain (86.1%), and had confusion or ‘brain fog’ (94.5%), for example (**Table 1**).

As we reported previously (2), about 63% of DecodeME cases indicated that they had an infection when, or just before, their ME/CFS symptoms first started (**Table 1**): glandular fever (infectious mononucleosis, 16.3%), the COVID-19 virus (SARS-CoV-2, 3.3%) or another infection (43.1%). ME/CFS symptoms were described by 17.2% as worsening, and by 71.7% as either fluctuating, or relapsing and remitting (**Table 1**).

### Genome-wide significant ME/CFS loci

Among the DecodeME DNA cohort of 16,730 participants, we inferred 15,579 (93.1%) to have European genetic ancestries. We then used REGENIE to compare imputed genotypes of cases against sex– and ancestry-matched UKB controls at 8,835,520 variants. As required for GWAS, there was good ancestry matching between the cases and controls and the PCs we used did not capture long-range LD structure (**Fig. S1, S2, S3**).

**Fig. 2:**
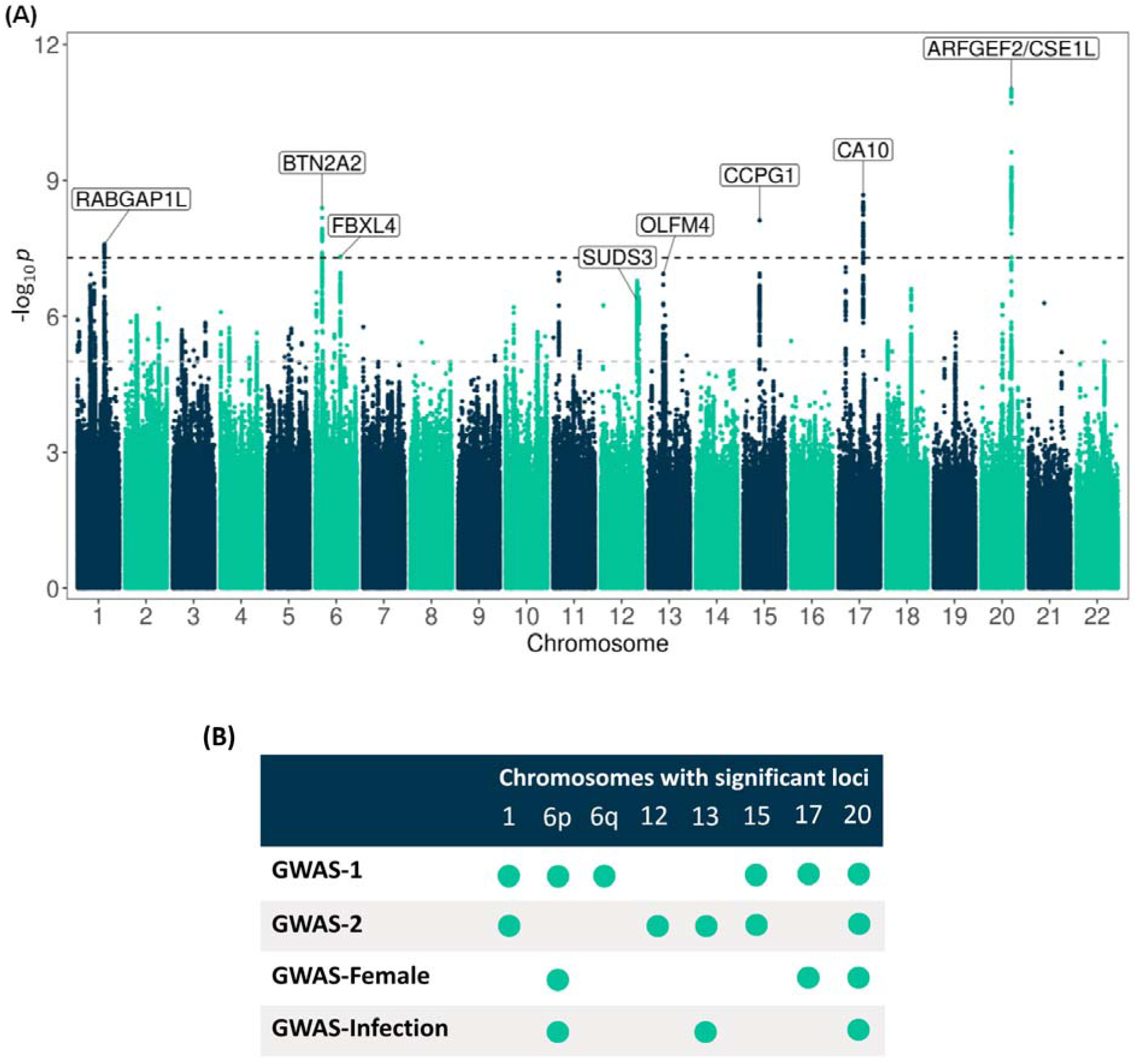
Eight genome-wide significant associations to ME/CFS. (A) Manhattan plot of the primary DecodeME genome-wide association study (GWAS-1), showing the chromosomal position of DNA variants (x-axis) against −log_10_p (*y*-axis) from genome-wide association tests accounting for sex and genetic ancestry. The upper horizontal dashed line indicates the threshold used to define genome-wide significant associations (*p*⍰<⍰5⍰×⍰10^−8^); the lower horizontal dashed line indicates p < 1 x 10^−5^. Six loci were genome-wide significant in GWAS-1. An additional locus, *OLFM4*, was genome-wide significant in a GWAS of cases reporting an infection prior to their ME/CFS symptoms. Prioritised genes are indicated. (B) Eight significant associations (*p*⍰<⍰5⍰×⍰10^−8^) were identified in GWAS-1 or GWAS-2 (which used all DecodeME cases), or in stratified comparisons with controls using only female cases (GWAS-Female) or cases reporting an infection prior to onset of ME/CFS symptoms (GWAS-Infection). Two lead variants were associated with ME/CFS in both chr6p and chr20 intervals for GWAS-1 and GWAS-Female. We found no associations for GWAS-Male or GWAS-No-Infection.

**Fig. 3.**
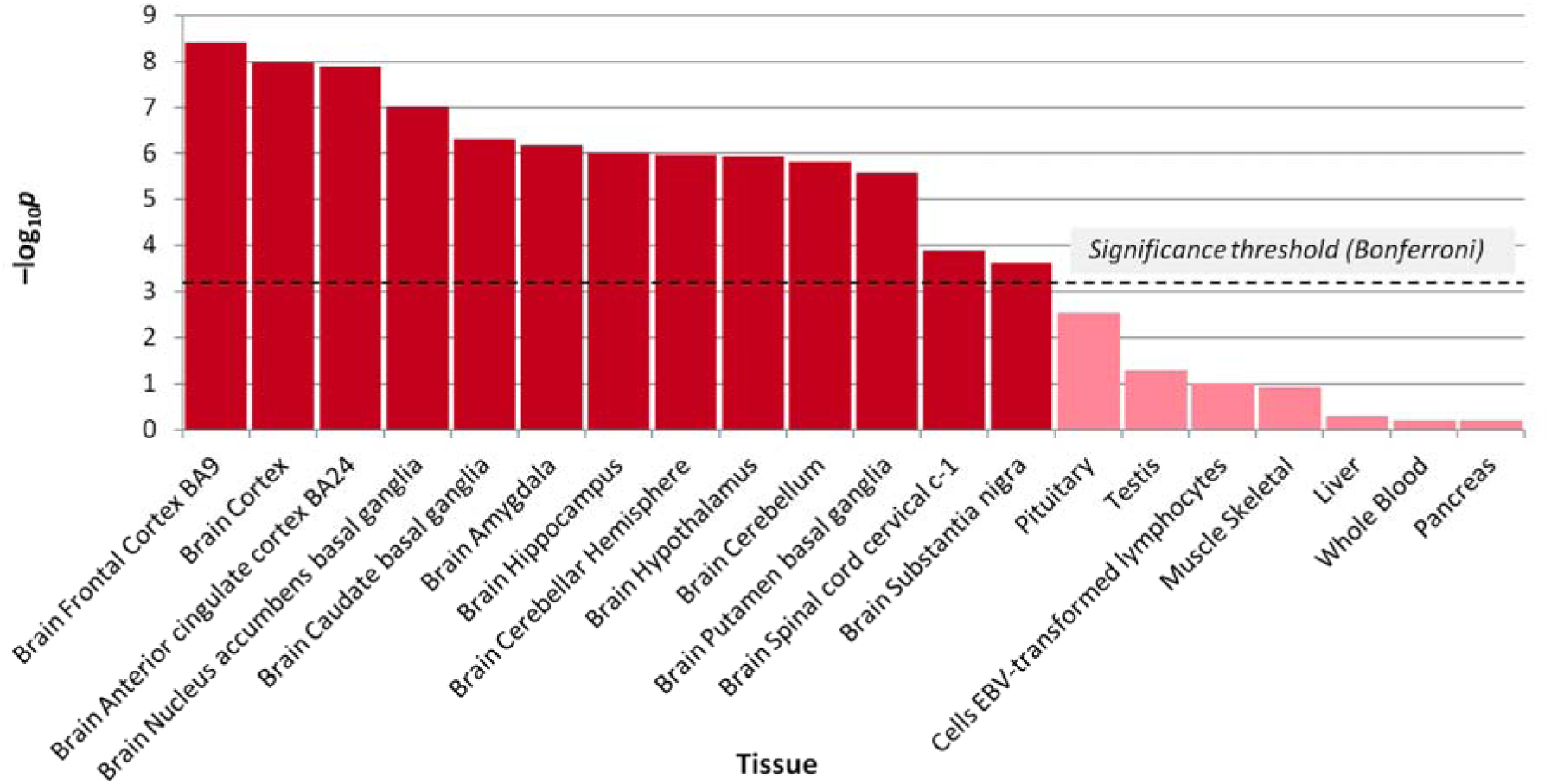
MAGMA gene-tissue analysis shows statistically significant enrichment of ME/CFS-related genes in all 13 brain tissues. *P*-values for 41 less significantly associated tissues are not shown. The significance threshold is *p* < 9.3×10^−4^ (= 0.05/54), 1-sided test.

We performed six GWAS involving DecodeME cases (**Table 2**), with matched European genetic ancestry (ongoing analyses are including all ancestries). The primary GWAS (GWAS-1) compared all DecodeME cases with the largest possible number of UKB controls to maximise discovery power. We did four additional GWAS that stratified by sex or by infection at disease onset. We also performed a further GWAS (GWAS-2) comparing all DecodeME cases with a limited set of UKB controls, as a comparison GWAS for independent UKB replication that used the rest of the UKB controls (described below: see **Replication**).

**Table 2.** Case and control groups in the six DecodeME GWAS.

| GWAS | Case group | n | Control group | n |
| --- | --- | --- | --- | --- |
| GWAS-1 | All DecodeME cases | 15,579 | All UKB controls | 259,909 |
| GWAS-Female | DecodeME female cases | 12,833 | UKB female controls | 218,949 |
| GWAS-Male | DecodeME male cases | 2,746 | UKB male controls | 40,960 |
| GWAS-No-Infection | DecodeME no-infection cases | 5,841 | All UKB controls | 259,909 |
| GWAS-Infection | DecodeME infection cases | 9,738 | All UKB controls | 259,909 |
| GWAS-2 | All DecodeME cases | 15,579 | UKB controls subset not used in UKB case-replications | 155,790 |

#### DecodeME GWAS: Overall and stratified analyses

Our primary GWAS, GWAS-1, compared 15,579 DecodeME cases with 259,909 UKB controls (case:control ratio of 1:17), across all autosomes. It yielded six genome-wide significant loci (*p* < 5 x 10^−8^; **Figs. 2A, 2B**; **Table 3**; **Table S1**; **Fig. S4**). Two of these loci (chr6p22.2 and chr20q13.13) each contained a pair of variants that were not in LD (*r*^2^ < 0.1) and that were both associated with ME/CFS. We estimated ME/CFS SNP-based heritability from GWAS-1, based on the LDSC method and reported on a liability scale. It was modest but significantly different from zero, with *h_l_*^2^ = 0.095 (SD = 0.006).

**Table 3.** Eight genome-wide significant ME/CFS loci (GRCh38 coordinates) in six DecodeME genome-wide association studies (GWAS). The most significant variant in GWAS-1 represents each locus (shown as chromosome: location: not-effect-allele: effect-allele). Lead variants in all GWAS are listed in **Table S1**. Case and control numbers for each GWAS are given in Table 2. Linked variants in chr6p22.2 and chr20q13.13 (6:26329103:A:T and 20:47529913:C:T) – not shown here – were genome-wide significant in the ‘Infection at onset’ GWAS, GWAS-Infection. A1FREQ is the frequency of the effect allele in the combined cohort. OR is the Odds-Ratio of ME/CFS risk in cases versus controls (with 95% confidence interval, [95% CI]). Lead genes are those discussed in the main text. –Log_10_*p* is the negative log_10_ of the test’s *p*-value, the statistical significance of association to a variant. Genome-wide significant values (*p* < 5 x 10^−8^) are shown in bold.

| GRCh38 |  | GWAS-1 |  |  | GWAS-Female |  |  | GWAS-Male |  |  |
| --- | --- | --- | --- | --- | --- | --- | --- | --- | --- | --- |
| SNPid | Prioritised genes | A1FREQ | OR (95% CI) | $-\log_{10}p$ | A1FREQ | OR (95% CI) | $-\log_{10}p$ | A1FREQ | OR (95% CI) | $-\log_{10}p$ |
| 1:173846152:T:C | *chr1q25.1* *RABGAP1L* | 0.325 | 0.927 (0.903 - 0.952) | **7.59** | 0.325 | 0.930 (0.903 - 0.958) | 5.93 | 0.325 | 0.892 (0.837 - 0.950) | 3.40 |
| 6:26239176:A:G | *chr6p22.2* *BTN2A2* | 0.261 | 1.086 (1.057 - 1.116) | **8.39** | 0.262 | 1.090 (1.058 - 1.124) | **7.75** | 0.258 | 1.069 (1.001 - 1.142) | 1.34 |
| 6:97984426:C:CA | *chr6q16.1* *FBXL4* | 0.546 | 0.934 (0.911 - 0.957) | **7.31** | 0.546 | 0.937 (0.912 - 0.963) | 5.64 | 0.548 | 0.920 (0.868 - 0.975) | 2.32 |
| 12:118202773:C(T^13):C | *chr12q24.23* *SUDS3* | 0.139 | 1.100 (1.061 - 1.140) | 6.78 | 0.139 | 1.104 (1.061 - 1.148) | 6.14 | 0.139 | 1.059 (0.973 - 1.153) | 0.73 |
| 13:53194927:GT:G | *chr13q14.3* *OLFM4* | 0.287 | 1.077 (1.048 - 1.107) | 6.94 | 0.287 | 1.080 (1.048 - 1.113) | 6.32 | 0.287 | 1.040 (0.974 - 1.110) | 0.62 |
| 15:54866724:A:G | *chr15q21.3* *CCPG1* | 0.312 | 1.082 (1.053 - 1.111) | **8.12** | 0.312 | 1.070 (1.039 - 1.102) | 5.23 | 0.310 | 1.146 (1.077 - 1.219) | 4.73 |
| 17:52183006:C:T | *chr17q22* *CA10* | 0.330 | 1.084 (1.056 - 1.113) | **8.67** | 0.329 | 1.088 (1.057 - 1.120) | **8.02** | 0.332 | 1.085 (1.020 - 1.155) | 2.00 |
| 20:48914387:T:TA | *chr20q13.13* *ARFGEF2/CSE1L* | 0.634 | 1.095 (1.067 - 1.124) | **11.02** | 0.633 | 1.097 (1.066 - 1.128) | **9.58** | 0.637 | 1.103 (1.037 - 1.173) | 2.72 |

| GRCh38 |  | GWAS-Infection |  |  | GWAS-No-Infection |  |  | GWAS-2 |  |  |
| --- | --- | --- | --- | --- | --- | --- | --- | --- | --- | --- |
| SNPid | Prioritised genes | A1FREQ | OR (95% CI) | $-\log_{10}p$ | A1FREQ | OR (95% CI) | $-\log_{10}p$ | A1FREQ | OR (95% CI) | $-\log_{10}p$ |
| 1:173846152:T:C | *chr1q25.1* *RABGAP1L* | 0.326 | 0.939 (0.909 - 0.970) | 3.79 | 0.326 | 0.899 (0.863 - 0.937) | 6.41 | 0.326 | 0.921 (0.896 - 0.947) | **8.18** |
| 6:26239176:A:G | *chr6p22.2* *BTN2A2* | 0.261 | 1.086 (1.050 - 1.123) | 5.77 | 0.261 | 1.083 (1.038 - 1.130) | 3.68 | 0.262 | 1.080 (1.050 - 1.112) | 6.95 |
| 6:97984426:C:CA | *chr6q16.1* *FBXL4* | 0.547 | 0.923 (0.895 - 0.951) | 6.82 | 0.547 | 0.948 (0.913 - 0.984) | 2.26 | 0.546 | 0.936 (0.912 - 0.960) | 6.50 |
| 12:118202773:C(T^13):C | *chr12q24.23* *SUDS3* | 0.139 | 1.092 (1.045 - 1.141) | 4.11 | 0.138 | 1.115 (1.056 - 1.177) | 4.09 | 0.139 | 1.111 (1.070 - 1.153) | **7.51** |
| 13:53194927:GT:G | *chr13q14.3* *OLFM4* | 0.287 | 1.100 (1.064 - 1.137) | **7.59** | 0.286 | 1.034 (0.992 - 1.079) | 0.93 | 0.287 | 1.082 (1.052 - 1.114) | 7.28 |
| 15:54866724:A:G | *chr15q21.3* *CCPG1* | 0.311 | 1.089 (1.054 - 1.125) | 6.52 | 0.311 | 1.072 (1.029 - 1.116) | 3.08 | 0.312 | 1.087 (1.058 - 1.118) | **8.46** |
| 17:52183006:C:T | *chr17q22* *CA10* | 0.329 | 1.076 (1.042 - 1.112) | 5.11 | 0.329 | 1.102 (1.058 - 1.147) | 5.60 | 0.330 | 1.079 (1.049 - 1.109) | 7.21 |
| 20:48914387:T:TA | *chr20q13.13* *ARFGEF2/CSE1L* | 0.633 | 1.092 (1.058 - 1.127) | 7.18 | 0.633 | 1.101 (1.058 - 1.146) | 5.67 | 0.635 | 1.100 (1.070 - 1.130) | **11.15** |

Two loci, *OLFM4* (chr13q14.3), and *TAOK3/SUDS3* (chr12q24.23), had not been genome-wide significant in GWAS-1 but were significant in GWAS-2. This brought the total number of ME/CFS-associated loci to eight (**Fig. 2B**; **Table 3**). We show LocusZoom representations of these loci in **Fig. S5**.

#### Females and males

In GWAS-Females, we tested females only (12,833 cases and 218,949 controls). This GWAS rediscovered three of the six significant loci found in GWAS-1 (**Fig. 2B**; **Table 3**). We identified no genome-wide significant associations when we tested males only (2,746 cases and 40,960 controls). Nevertheless, two of the three variants that were genome-wide significant in females were replicated in males (chr20q13.13 and chr17q22) at a Bonferroni-corrected significance of *p* < 0.05/3 (**Table 3**).

The study’s large numbers of ME/CFS cases allowed us to test for genetic differences in ME/CFS risk between females and males using a two-tailed Student’s t-test (40). None of the eight DecodeME genome-wide significant loci yielded evidence for sex bias (*p* < 0.05; **Table S2**). This is also seen in the overlapping confidence intervals of effect estimates between males and females (**Table 3**).

GWAS-2 analysed all 15,579 DecodeME cases and 155,790 matched UKB controls (fewer controls than in GWAS-1). It yielded five genome-wide significant associations (**Table S1**). They were the three that were significant in GWAS-1, the additional chr13q14.3 association found in GWAS-Infection, and a new association within chr12q24.23 (TAOK3/SUDS3) that in GWAS-1 had fallen just below the significance threshold (*p* = 1.6 x 10^−7^).

#### Infection and non-infection onset

Next, we conducted GWAS-Infection, a GWAS with cases who reported an infection when, or just before, their first ME/CFS symptoms started (9,738 cases and 259,909 controls). It yielded three genome-wide significant loci. These overlapped two of the initial six found in GWAS-1 (chr6p22.2 and chr20q13.13; **Fig. 2B**; **Table S1**). The third significant association in GWAS-Infection lay near *OLFM4* (chr13q14.3). This association was stronger in GWAS-Infection (β = 0.095 ± 0.017; –log_10_*p* = 7.59) than in GWAS-No-Infection (β = 0.034 ± 0.022; –log_10_*p* = 0.93), and nearly reached genome-wide significance in GWAS-1 (**Table 3**).

We also tested for associations using only cases who did not report infection at onset (5,841 cases and 259,909 controls; ‘GWAS-No-Infection’). This yielded no significant associations (**Table 3**).

These six GWAS above showed little evidence of genomic inflation (λ = 1.003–1.066), which supports these results being robust to residual population stratification.

### Replication

We sought to replicate DecodeME’s genome-wide significant associations in cases from groups of biobanks (**Table 4**). We looked first at cases defined to include post-exertional malaise – the cardinal symptom of ME/CFS – and then more broadly defined to not require post-exertional malaise. This was because more accurate case definition and larger numbers of cases could each give more power to detect genetic associations.

**Table 4.** Case and control groups in the four external replication analyses.

| GWAS | Case group | n | Control group | n |
| --- | --- | --- | --- | --- |
| R-1 | Lifelines and UKB Biobank cases: required post-exertional malaise-like symptoms and fatigue | 13,767 | Controls drawn from the same biobank | 212,183 |
| R-2 | Seven other cohorts' cases: did not require post-exertional malaise and fatigue | 15,251 | Controls drawn from the same cohort | 1,878,066 |

#### UK Biobank

For our UKB replication attempt, we plan two GWAS using cases and controls from the UKB, and created GWAS-2, a new DecodeME GWAS (described above), as an appropriate comparison for them. This design also makes it appropriate to combine DecodeME GWAS-2 results with UKB ME/CFS in future meta-analysis. Results of the replication UKB GWAS will be provided in due course.

#### External replication analyses

We attempted to replicate DecodeME findings with two analyses using cases defined in European and US biobanks (**Table 4**). The first, R-1 had 13,767 cases and 212,183 controls from Lifelines and UKB. Lifelines cases were clinically diagnosed with ME/CFS and had post-exertional malaise. UKB cases had persistent tiredness that remained after sleep, and were tired after minimal physical or mental exertion, but were not necessarily diagnosed with ME/CFS (**Supplementary Methods**). The R-1 GWAS showed no evidence of genomic inflation (λ =1.007). R-2 (14,250 cases and 1,433,490 controls from six national cohorts) defined cases using only electronic health records (ICD10:G93.3 or Phecode:Phe_798_1, or related SNOMED code), without any requirement for post-exertional malaise or fatigue (**Supplementary Methods**).

In R-1, we tested for replication 22 of the 25 most significant DecodeME GWAS-1 associations, defined at a less stringent threshold of *p* ≤ 9.4 x 10^−7^. We could not test the three other associations because neither markers nor suitable proxies were available in the results of these analyses. No replicated associations were identified in R-1 after correcting for multiple hypothesis testing (*p* < 0.05/22; **Table S3**), and similarly none in R-2 for the 23 tests that could be performed (*p* < 0.05/23; **Table S3**). However, 9 of the 22 GWAS-1 associations were associated in R-1 with *p*-values < 0.05, a larger proportion than expected by chance. These nine included four DecodeME loci (*RABGAP1L, FBXL4, OLFM4, CA10*), plus *LRRC7* – a gene associated by MAGMA gene-based testing (below, **Table S4**) – and *DCC*, a gene that has repeatedly been associated with chronic pain (41).

### MAGMA analysis

Next, we tested for positive relationships between gene expression in a tissue type and gene-based ME/CFS association strengths, using MAGMA (42). Thirteen genes were significantly associated with ME/CFS in a MAGMA gene-based test of 18,637 genes (*p* < 0.05/18637; **Table S4**). We considered 54 tissue types and identified significant enrichment of these genes’ expression for 13 (*p* < 0.05/54), all of which were brain regions (**Fig. 3**). MAGMA analysis found no significant associations between other gene sets and ME/CFS after applying the Bonferroni correction for multiple tests (*p_Bonferroni_* < 0.05).

### Gene prioritisation

Linking GWAS variants to causal genes that may provide biological insights and medical applications remains a challenge for the field (43). There were 43 protein-coding genes with at least one eQTL within an ME/CFS genome-wide significant interval, and we prioritised 29 ME/CFS candidate causal genes among them (**Fig. 4**; **Table S5**; **Table S6**; **Fig. S6**). These ‘Tier 1’ genes have a high (≥75%) posterior probability for colocalisation (H4) of a shared causal variant for their expression and ME/CFS risk in at least one of 50 tissues. We calculated this probability using *coloc*, a statistical method that predicts whether two traits are likely to be influenced by the same genetic variant in a specific chromosomal region (32). For this step, we disregarded the histone genes in the chr6p22.2 HIST1 cluster as candidates, as their sequences and functions are highly redundant (44).

**Fig. 4:**
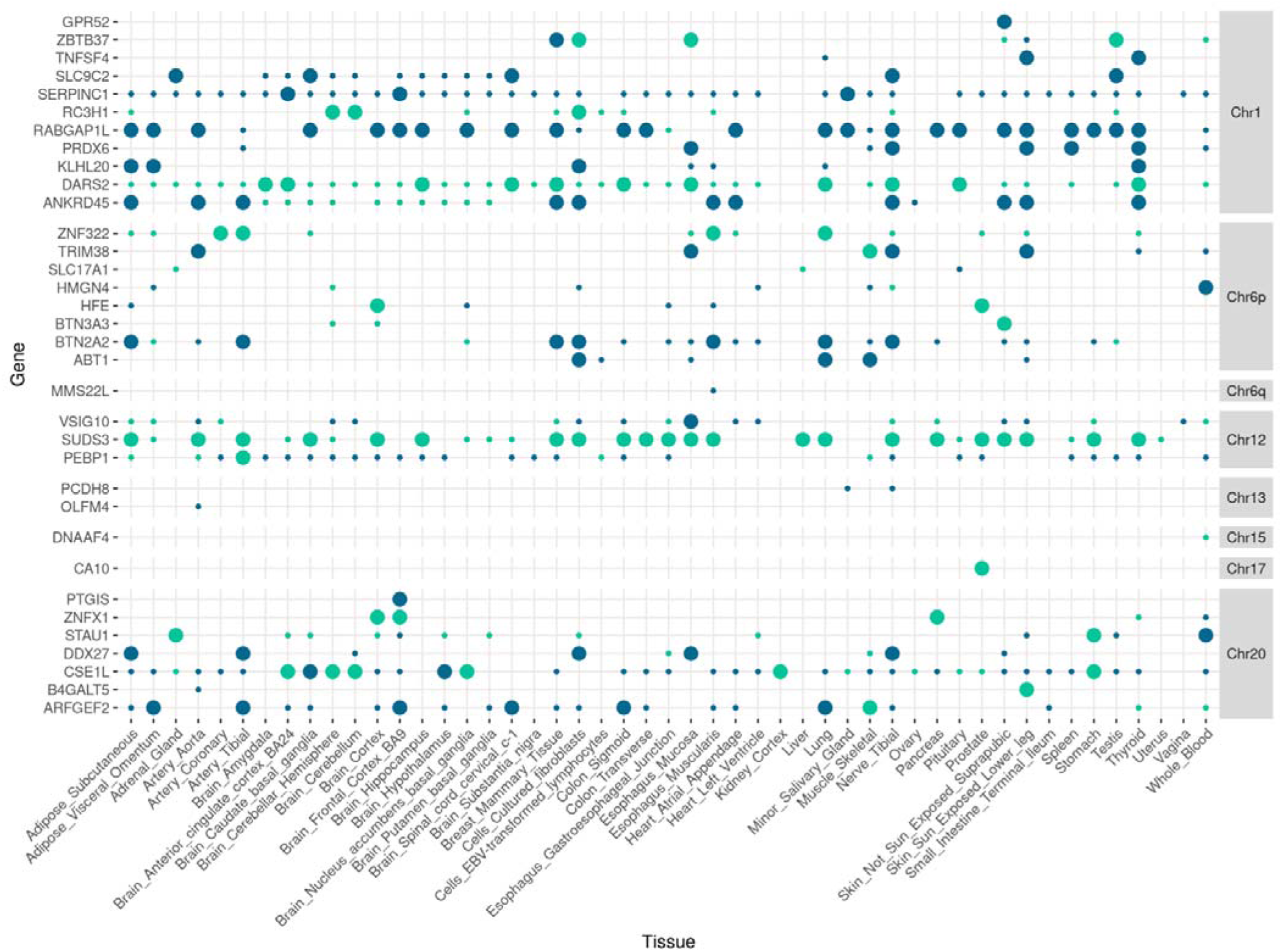
Approximate Bayes factor posterior probability (PPH4) that mRNA expression and ME/CFS traits are associated and share a single causal variant. Thirty-four protein coding genes (*y*-axis) with at least one expression QTL (eQTL; GTEx-v10) within a FUMA-defined ME/CFS interval that show colocalisation with the eQTLs for at least one of 49 GTEx-v10 tissues (*x*-axis). Green circles indicate that increased ME/CFS risk allele is associated with increased gene expression for eQTLs in the FUMA-defined interval. Blue circles indicate that increased ME/CFS risk allele is associated with decreased gene expression for eQTLs in the FUMA-defined interval, and vice-versa. The area of each circle indicates the test’s value of PPH4 (posterior probability for single shared causal variant): smaller circles indicate 0 < PPH4 < 0.75, and larger ones indicate PPH4 ≥ 0.75.

#### Tier 1 genes

The chr1q25.1 interval is associated with 11 Tier 1 genes (**Table S5**). Of these, *RABGAP1L* is notable for its eQTLs colocalising with ME/CFS risk in 24 of the 50 tissue samples (**Fig. 4**). RABGAP1L promotes expulsion of the bacterium *Streptococcus pyogenes* from cells via endocytosis (45) and also limits replication of multiple viruses (46). The ME/CFS risk increasing allele is associated with decreasing *RABGAP1L* gene expression for GTEx-v10 eQTLs in the FUMA-defined interval, and vice-versa (**Fig. 4**). This suggests that ME/CFS-associated *RABGAP1L* variants could enhance susceptibility to bacterial and viral infection.

The chr6p22.2 locus includes seven Tier 1 genes, with *BTN2A2* and *TRIM38* eQTLs colocalised with ME/CFS risk in the most tissues (7 and 5, respectively). Complexes involving butyrophilin-3 and –2 homologues (*BTN3A1*, *BTN3A2*, *BTN3A3*, *BTN2A1* and *BTN2A2*) allow innate-like Vγ9Vδ2 T cells to distinguish self-derived from non-self-derived pyrophosphate antigens (pAgs) (47). Mouse *Btn2a2* knockout model phenotypes (48,49) – together with the discordance of *BTN2A2* expression and ME/CFS risk effects (**Fig. 4**) – imply that the ME/CFS risk-increasing allele could impair T-cell responses and worsen autoimmune disease. TRIM38 is known to prevent an excessive response to DNA viral infection and uncontrolled inflammation (50). Depending on tissue, the ME/CFS risk-increasing allele is associated with decreased or increased *TRIM38* expression (**Fig. 4**).

The chr12q24.23 ME/CFS-associated locus contains three Tier 1 genes, including *SUDS3*, whose genetic expression signal is significantly colocalised and concordant with ME/CFS risk in 22 GTEx-v10 tissues (**Fig. 4**). *SUDS3* encodes a protein that is a negative regulator of microglial inflammation (51). ME/CFS genetic risk at this locus, therefore, could act to suppress the microglial inflammatory response.

The only Tier 1 gene in the chr17q22 interval was *CA10*, whose genetic variation could begin to explain the pain experienced by over 86% of DecodeME’s ME/CFS cases (**Table 1**). The CA10 protein inhibits the addition of heparan sulfate glycosaminoglycan to presynaptic neurexins. This post-translation modification enables their binding to postsynaptic neuroligins (52,53). This trans-synaptic interaction is critical for synaptic transmission and, when disrupted in a rodent model, prevents pain (54).

Of seven Tier 1 genes in chr20q13.13, eQTLs for *CSE1L* and *ARFGEF2* colocalised with ME/CFS risk in 8 and 7 tissues, respectively. CSE1L helps to re-export importin-alpha from the nucleus to the cytoplasm after it releases its import cargos into the nucleoplasm (55). A small molecule inhibitor of CSE1L reduces the nuclear translocation of transcriptional factor SP1 in macrophages, leading to decreased TNF-α release, a marker of suppressed inflammation (56). ARFGEF2 is involved in releasing the TNF-α receptor in exosome-like vesicles (57). A third candidate gene in this associated interval is *ZNFX1*, an interferon-stimulated dsRNA sensor that restricts the replication of RNA viruses early after infection (58).

#### Tier 2 genes

It might not have been possible to find Tier 1 genes in intervals associated with our significant SNPs because available eQTL resources are incomplete, some eQTL are only detectable upon stimulation of cells (59), or because the single causal variant assumption we used does not always hold. For intervals without Tier 1 genes, we defined Tier 2 genes as the closest protein-coding genes to the lead variant: this yielded *FBXL4* (chr6q16.1), *OLFM4* (chr13q14.3), and *CCPG1* (chr15q21.3). Mutations in *FBXL4* cause increased mitophagy, and mitochondrial DNA depletion (60). Olfactomedin-4 (OLFM4) suppresses antibacterial and inflammatory responses by binding to neutrophil cationic proteins and neutralises their ability to kill bacteria and form immunogenic complexes with DNA (61). CCPG1 mediates the selective degradation of the endoplasmic reticulum by autophagy (62). This is a host defense mechanism when pathogens infect cells, and its deficiency facilitates viral infection (63).

### Human leukocyte antigen alleles

We tested for association of ME/CFS status to HLA alleles. For this, we imputed alleles in cases following the imputation approach used for UKB controls (25). A single class II HLA allele (HLA-DQA1*05:01) was associated with ME/CFS at genome-wide significance (*p* = 1.4 x 10^−10^; **Fig. S7**; **Table S7**). The frequency of HLA-DQA1*05:01 among cases (21.7%) was lower than among controls (23.2%) and so is predicted to protect against ME/CFS. This association was robust to testing restricted to the genetically more homogeneous White British genetic ancestries subset (**Supplementary Methods**; *p* = 7.2 x 10^−8^). Nevertheless, we did not find associations to HLA-DRB1*03:01 (*p*=0.27) or HLA-DQB1*02:01 (*p*=0.042), despite expecting them due to their strong linkage disequilibrium with HLA-DQA1*05:01. We will need to repeat this analysis using HLA alleles that have been imputed for cases and controls jointly in order to verify (or dismiss) the potential association.

### Shared associations with other traits

Three out of our eight ME/CFS-associated intervals had previously been associated to depression (chr1q25.1, chr13q14.3 and chr20q13.13) (64,65), and one locus to pain (chr17q22) (41) phenotypes. Where these studies provided full summary statistics, we used *coloc* (32) to investigate the level of support for these genetic signals and our ME/CFS results being underpinned by the same causal variant.

The ME/CFS and depression phenotype genetic signals did not colocalise at the three loci investigated (**Fig. 5**; **Table S8**). For a region within chr17q22 (**Fig. 5E**), which we subdivided due to its complex association structure (**Fig. 5F**), the ME/CFS and multisite chronic pain associations showed significant colocalisation (approximate Bayes factor posterior probability for shared causal variant, 92.8%). In summary, we found no evidence for the same genetic variants causing both ME/CFS and depression phenotypes within the loci tested, but we did find significant evidence for a shared genetic signal explaining ME/CFS and multisite chronic pain phenotypes at chr17q22 (*CA10*).

**Fig. 5.**
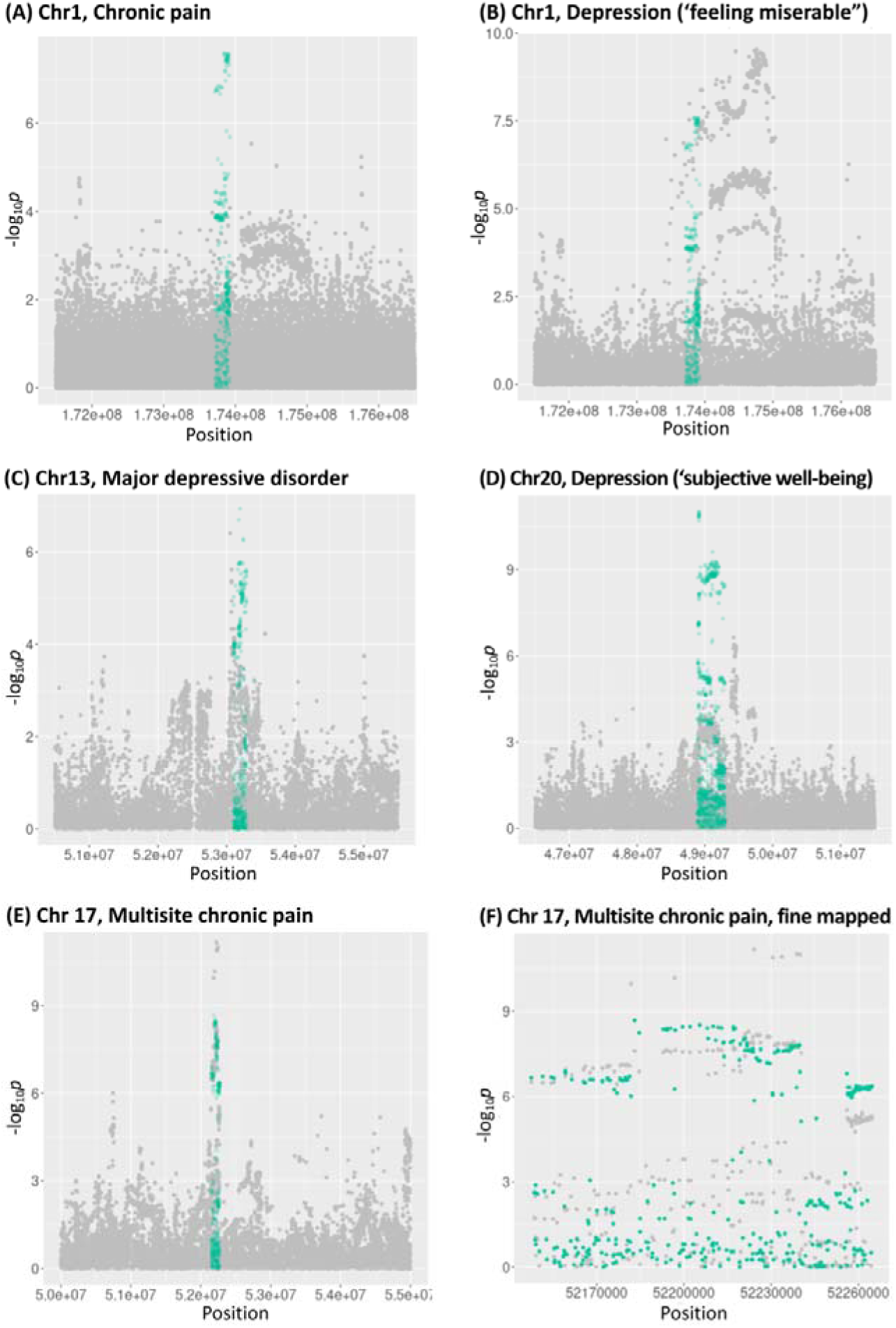
Regional association plots for ME/CFS and non-ME/CFS traits. Non-ME/CFS data points are shown in grey and ME/CFS data points for the FUMA-defined interval only are shown in green. The *x*-axis is the chromosomal position. The *y*-axis is –log_10_*p* of the association. (A) chr1q25.1 for chronic pain. (B) chr1q25.1 for depression (“feeling miserable”). (C) chr13q14.3 for major depressive disorder. (D) chr20q13.13 for depression (“subjective well-being”). (E) chr17q22 for multisite chronic pain, with (F) showing a subdivision of (E).

Two GWAS (66,67) reported associations to anxiety near *BTN2A2*. We could not perform *coloc* analysis because their full summary statistics are not available. However, low LD between lead variants (*R^2^* < 0.05 in the British in England and Scotland [GBR] population) indicate that the genetic signals contributing to ME/CFS and anxiety at the chr6p22.2 locus are distinct.

## DISCUSSION

DecodeME sought to identify common genetic variants linked with ME/CFS, and succeeded in finding a total of eight significant associations (**Fig. 2B**; **Table 3**).

DecodeME’s results, grounded in the principles of statistical genetics, now place ME/CFS research on a firm biological foundation: they begin to explain the disease’s heritable component (12,13), they improve the likelihood of finding effective drugs for ME/CFS (68), and they place this long-neglected disease on more equal terms with other common genetic conditions (69). DecodeME further provides a much-needed managed resource of genotype and phenotype data. It also offers researchers a pool of thousands of DecodeME participants who consented to be recontacted to take part in future epidemiological, genetic, and biomolecular studies, including clinical trials for diagnostics and therapeutics.

DecodeME’s success is likely to be because of its high statistical power, due both to our recruitment methods and use of well-defined case criteria. Direct recruitment to DecodeME lowered the barriers to engagement, especially for those who are housebound or bedbound (**Supplementary Methods**), and DecodeME’s PPI (patient and public involvement) probably also greatly boosted recruitment by helping gain the trust of potential participants. Regarding case criteria, we used two international sets of criteria (5,26) and required cases to report a medical diagnosis and evidence for the key symptom of post-exertional malaise. These stringent criteria will have prevented the cohort being diluted by less clear-cut cases. The resulting cohort of 21,620 people who met case criteria makes DecodeME the largest ME/CFS study in the world to date.

Due to its large numbers of cases and controls, our study has substantially better statistical power to discover true associations than previous GWAS. Results from smaller-scale studies have not previously been replicated (16,21), including here by DecodeME. We achieved replication within our study: after accounting for multiple tests, two of the three signals that were significant genome-wide in females were also significant in males at α = 0.05. Also, for each of two loci, a pair of unlinked variants (*r*^2^ < 0.1) were separately associated with ME/CFS, indicating two causal signals.

Our results were not replicated in an analysis of data from 15,251 cases and 1,878,066 controls assembled across seven national biobanks (R-2). This could be due to chance, or differences in case definition or ascertainment bias. DecodeME’s ME/CFS case-selection was based on international criteria, and evidence for a clinical diagnosis and post-exertional malaise, ME/CFS’s hallmark symptom. Many cases for R-2 may have been given a clinical diagnosis of ME/CFS yet did not meet international case criteria (70), or had postviral fatigue syndrome without post-exertional malaise, or had chronic fatigue but not ME/CFS.

In another replication analysis (R-1), we compared 13,767 cases that were more narrowly defined and 212,183 controls, from two biobanks. This provided evidence of replication for 8 out of 21 of GWAS-1’s less significant associations, but only after applying a *p*-value threshold of 0.05 without correcting for multiple tests. Variation in how post-exertional malaise and ME/CFS diagnoses are recorded in clinical practice and biobanks could explain why replication was not stronger.

ME/CFS can be misdiagnosed as depression (71). The eight genome-wide significant associations for ME/CFS are different from genetic associations reported for depression. ME/CFS associations near to chr1q25.1, chr13q14.3 and chr20q13.13 were close to, but did not significantly colocalise with, signals for depression. However, near *CA10* (chr17q22), a region within the ME/CFS association did significantly colocalise to known chronic pain signals.

Long COVID, a condition with similar symptoms and starting, like most cases of ME/CFS, after an infection, has one genetic association near *FOXP4* (72), but this was not shared with ME/CFS (*p* = 0.59 for rs9367106).

Brain-expressed genes contributed to ME/CFS risk (**Fig. 4**). One such gene is *CA10*, whose shared association between pain and ME/CFS (**Fig. 5E**) may reflect disruption of *trans*-synaptic interactions between neurexins and neuroligins. ME/CFS-associated intervals also contain genes involved in innate immunity (*BTN2A2*, *OLFM4*, *RABGAP1L* and *ZNFX1*). These associations could shed light on why most people with ME/CFS report an infection when, or just before, their first ME/CFS symptoms started. *OLFM4’s* effect size is notably stronger among cases who reported an infection at onset than among those who did not. Drugs targeting these genes’ proteins might help protect against the consequences of microbial infection and therefore could reduce the risk of acquiring ME/CFS (72,73).

These DecodeME results have not shown why ME/CFS is a female-biased disease: none of its eight genetic signals show significant sex-bias. We have yet to test for association to chromosome X (or Y) variants.

We will need to carry out further analyses to replicate results, to fine-map candidate causal variants from DecodeME’s large number of variants with statistically significant associations, to show experimentally how, when and where they might exert their function, and to analyse DecodeME DNA data from all genetic ancestries. We will also further fine-map our eight associated regions.

Our study was limited to finding statistical associations between relatively common DNA variants (MAF ≥ 1%) and ME/CFS. Future whole genome sequencing of 18,824 DNA samples stored by DecodeME could reveal rare and structural variants that contribute to ME/CFS risk.

DecodeME has provided the first robust evidence that genetic variation contributes to the risk of developing ME/CFS, which should help to reduce the stigma of the illness. We are making full summary statistics and consented and pseudonymised individual-level data available via controlled access (73) to accelerate the search for effective treatments for this devastating condition (74).

### Ethics approval

The DecodeME study was reviewed and given a favourable opinion by the North West – Liverpool Central Research Ethics Committee (21/NW/0169) in June 2021 after we prepared and submitted our protocol, data analysis plan, data protection assessment, privacy policy and participant information document. The approval also included our participant invitation and reminders, and the consent-process text. The committee also approved a substantial amendment in October 2021. DecodeME is registered in the Research Registry (identifying number 6395).

## Acknowledgements

DecodeME would not have been possible without people with ME/CFS and their carers who gave their time and energy over many years. Current and past members of the UK CFS/ME Research Collaborative (now UK ME Research Collaborative) and its Patient Advisory Group had substantial input into the funding application and early stages of DecodeME. This research has been conducted using the UK Biobank Resource under Application Number 76173. This work uses data provided by patients and collected by the NHS as part of their care and support. The CureME team at the London School of Hygiene and Tropical Medicine contributed to the funded grant application. We are very grateful to Professor Sir Stephen Holgate and the DecodeME Scientific Advisory Board, which includes Professors Brian Hughes, Benedicte A. Lie, and Jonathan Edwards, for their critical contributions. We thank Zameel Cader, Caleb Webber, and Jimena Monzon Sandoval for providing a study’s summary statistics before its publication. We thank Simone Ribunacci for help and guidance with the UK Biobank imputation pipeline. We thank Audrey Ryback for helpful discussions.

## FinnGen

We want to acknowledge the participants and investigators of the FinnGen study (75).

## Lifelines

The Lifelines initiative has been made possible by subsidy from the Dutch Ministry of Health, Welfare and Sport, the Dutch Ministry of Economic Affairs, the University Medical Center Groningen (UMCG), Groningen University, and the Provinces in the North of the Netherlands (Drenthe, Friesland, Groningen).

## Genes & Health

We thank Social Action for Health, Centre of The Cell, members of our Community Advisory Group, and staff who have recruited and collected data from volunteers. We thank the NIHR National Biosample Centre (UK Biocentre), the Social Genetic & Developmental Psychiatry Centre (King’s College London), Wellcome Sanger Institute, and Broad Institute for sample processing, genotyping, sequencing and variant annotation. This work uses data provided by patients and collected by the NHS as part of their care and support. This research utilised Queen Mary University of London’s Apocrita HPC facility, supported by QMUL Research-IT, http://doi.org/10.5281/zenodo.438045

We thank: Barts Health NHS Trust, NHS Clinical Commissioning Groups (City and Hackney, Waltham Forest, Tower Hamlets, Newham, Redbridge, Havering, Barking and Dagenham), East London NHS Foundation Trust, Bradford Teaching Hospitals NHS Foundation Trust, Public Health England (especially David Wyllie), Discovery Data Service/Endeavour Health Charitable Trust (especially David Stables), Voror Health Technologies Ltd (especially Sophie Don), NHS England (for what was NHS Digital), for GDPR-compliant data-sharing backed by individual written informed consent.

A favourable ethical opinion for the main Genes & Health research study was granted by NRES Committee London – South East (reference 14/LO/1240) on 16 Sept 2014. Queen Mary University of London is the Sponsor, and Data Controller.

Most of all we thank all of the volunteers participating in Genes & Health.

We want to acknowledge the participants of the Estonian Biobank (EstBB) for their contributions. The Estonian Genome Center analyses were partly carried out in the High Performance Computing Center, University of Tartu. The activities of the EstBB are regulated by the Human Genes Research Act, which was adopted in 2000 specifically for the operations of EstBB. Individual-level data analysis in EstBB was carried out under ethical approval 1.1-12/624 from the Estonian Committee on Bioethics and Human Research (Estonian Ministry of Social Affairs), using data according to release application 6-7/GI/2015 from the Estonian Biobank.

We wish to thank the many participants in the Partners HealthCare Biobank (Mass General Brigham Biobank) for their willingness to engage in a research programme to advance our understanding of human health. We would also like to acknowledge all the recruiters and the participants of the Estonian Genome Center of the University of Tartu biobank. We thank MVP and the Michigan Genomics Initiative participants for their service and contributions to this study.

## Competing interests

ADC was until April 2025 a committee member of the Science for ME online support discussion forum.

CPP has had PhD studentships funded by Action for ME, the Chief Scientist Office (Scotland), and ME Research UK.

CS is the honorary medical adviser to the ME Association.

JW was on the UK ME/CFS Research Collaborative Patient Advisory Group and received travel expenses for participation in board meetings.

NM has authored an online e-learning module on ME/CFS and has received consultancy fees from the Learn about ME Project and Ono Pharmaceuticals as well as an honorarium from the GW4 ME/CFS Carers Project but has no direct or financial conflict of interest to declare in relation to this research.

SC is CEO of Action for ME; co-chair of the World ME Alliance; and Chair, Future Ambitions Board, B&NES.

SL is head of advocacy and communications of the World ME Alliance.

SMcG was formerly involved in the UK CFS/ME Research Collaborative Patient Advisory Group.

## Data availability

Supplementary Information, including the DecodeME questionnaire, is available from the Open Science Framework website: https://osf.io/rgqs3/. DecodeME anonymised data allowing investigation of this study’s consented data is available to researchers by managed access via our data access committee, see: <u>Data Access for Researchers</u>. This committee consists of a scientist, a person with ME/CFS and an ME/CFS charity representative, who strictly control access to the data. DecodeME’s anonymised and consented data are only shared with studies that meet high standards and whose academic or industrial researchers agree to treat its data with respect and to keep it secure. GWAS Summary Statistics are available for download from the DecodeME Summary Statistics folder of https://osf.io/rgqs3/files/osfstorage. For each GWAS, we provide a p-value, an effect size and its standard error, and the effect allele for each variant tested.

## Web links

UK Biobank: https://www.ukbiobank.ac.uk/

PLINK: https://www.cog-genomics.org/plink/

REGENIE: https://rgcgithub.github.io/regenie/overview/

UK Biobank project: https://www.ukbiobank.ac.uk/enable-your-research/approved-research/genome-wide-association-study-of-myalgic-encephalomyelitis-chronic-fatigue-syndrome-me-cfs

DecodeME Project Website.

## FIG

**Fig. S1:**
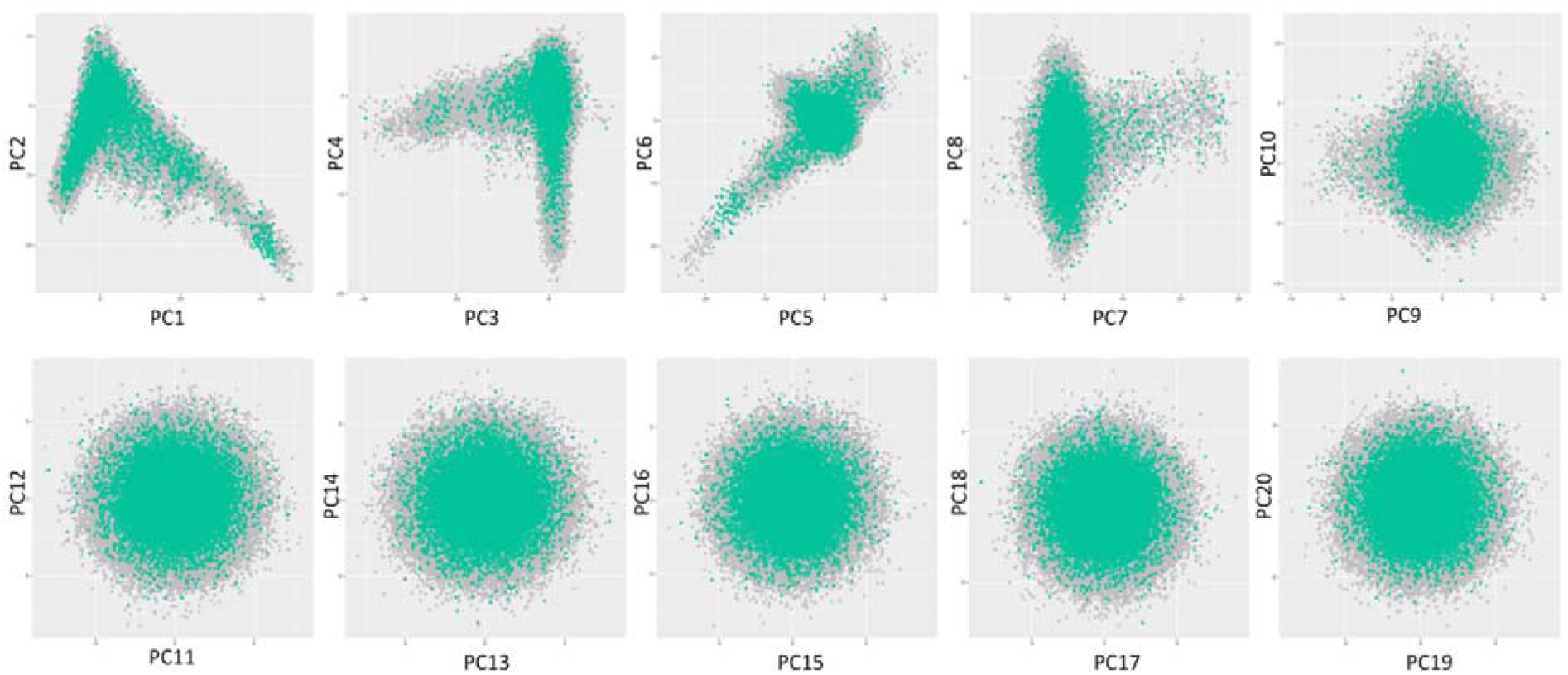
The principal component (PC) scores along each axis for PC1–20. Controls are shown in black, cases in green. After filtering, we noted that there is good matching between the cases and controls across all 20 PCs.

**Fig. S2:**
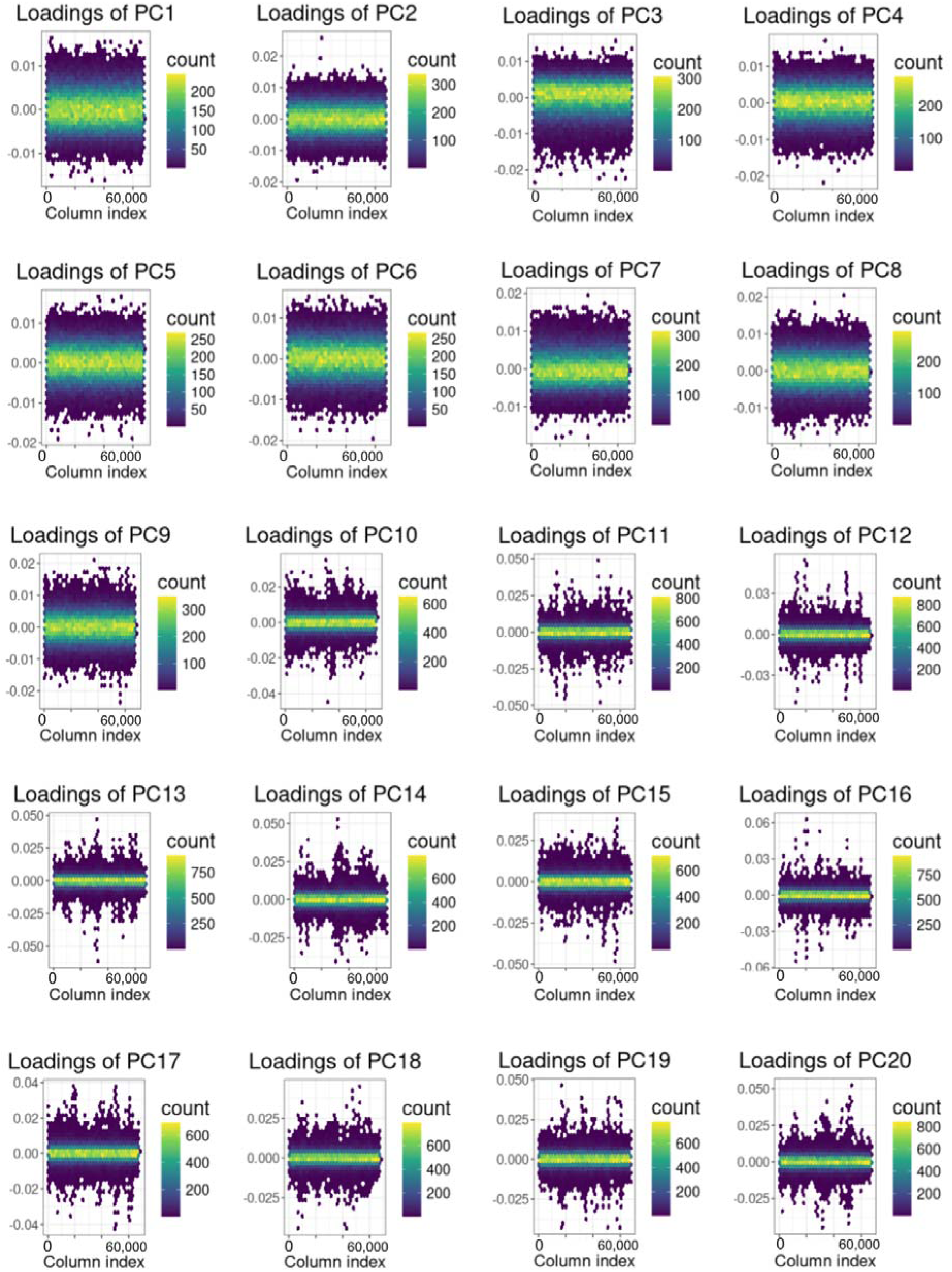
DNA variants’ loadings for principal components 1–20. We noted the absence of signal that might have indicated long-range linkage disequilibrium.

**Fig. S3:**
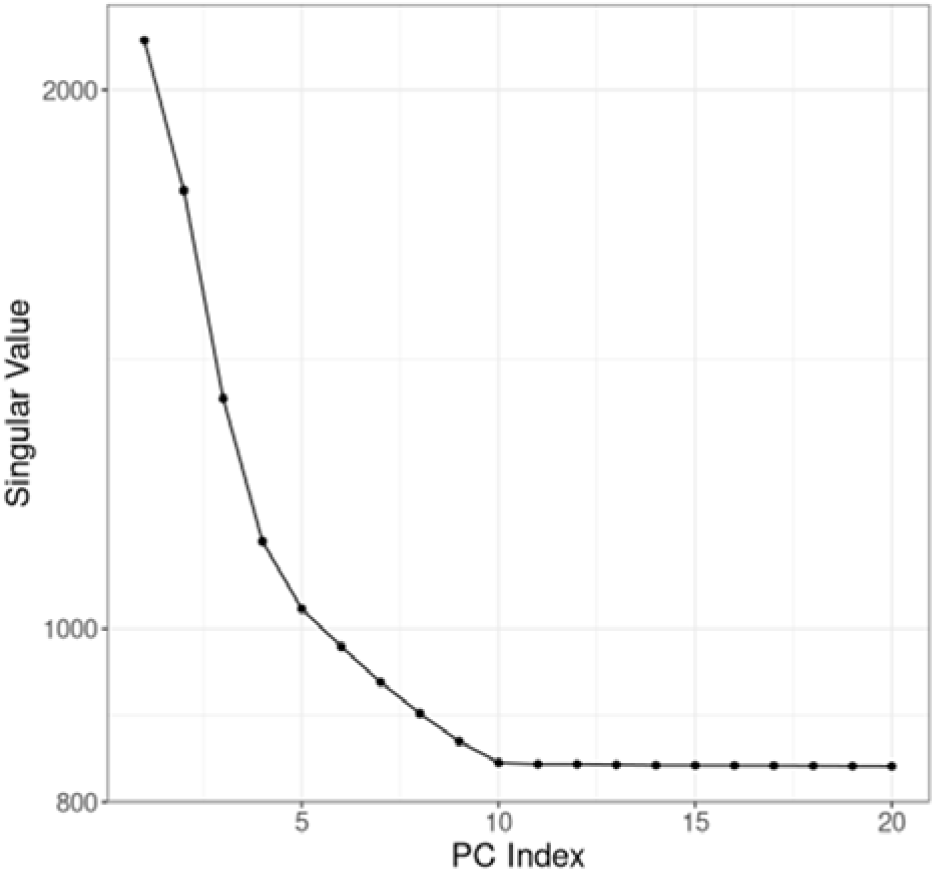
Scree plot indicating the relative contribution of each principal component (PC). The plot shows singular values from the singular value decomposition. We elected to use the first 20 PCs, a common choice in GWAS.

**Fig. S4:**
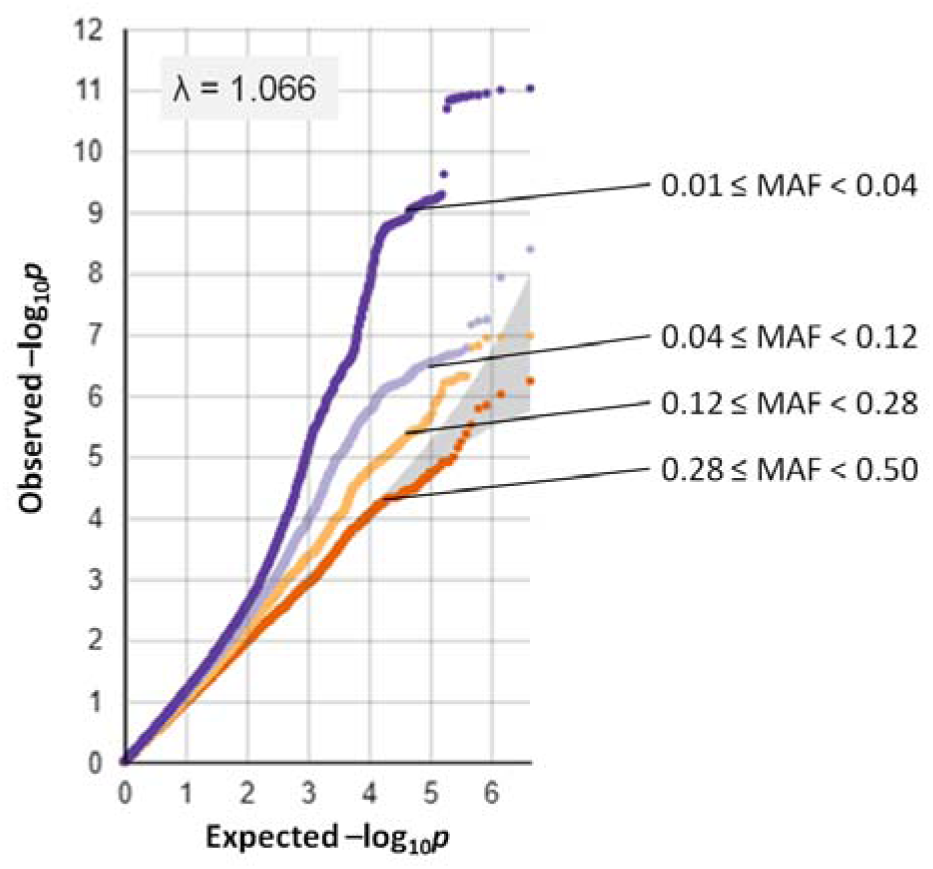
Quantile-Quantile (QQ) plots for GWAS-1. The plots show quantiles of observed association *p*-values (*y*-axis, on a –log_10_ scale) against the quantiles of the theoretical distribution for DNA variants binned by four different minor allele frequency (MAF) ranges. The genomic inflation factor calculated based on the 50th percentile was λ=1.066.

**Fig. S5:**
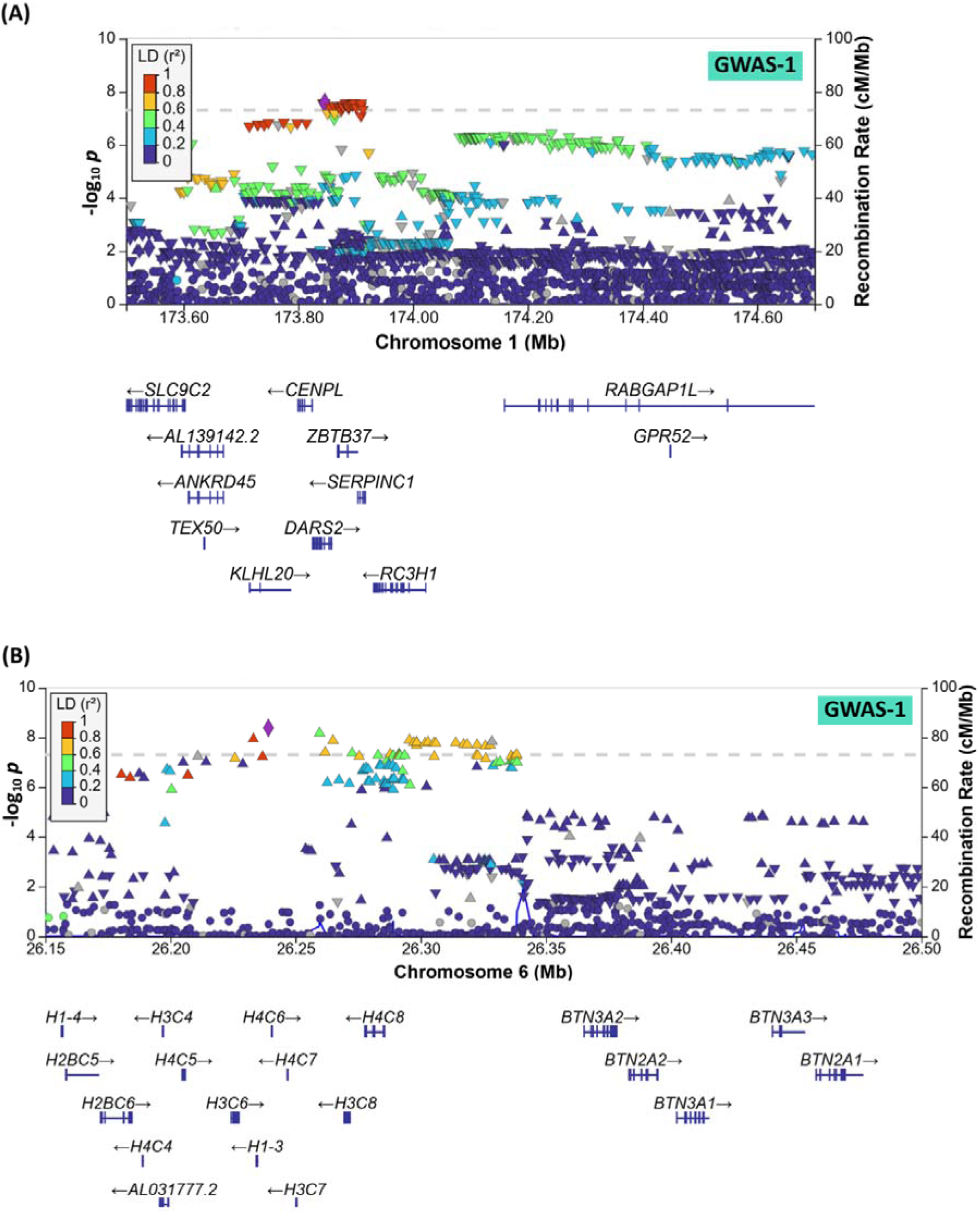
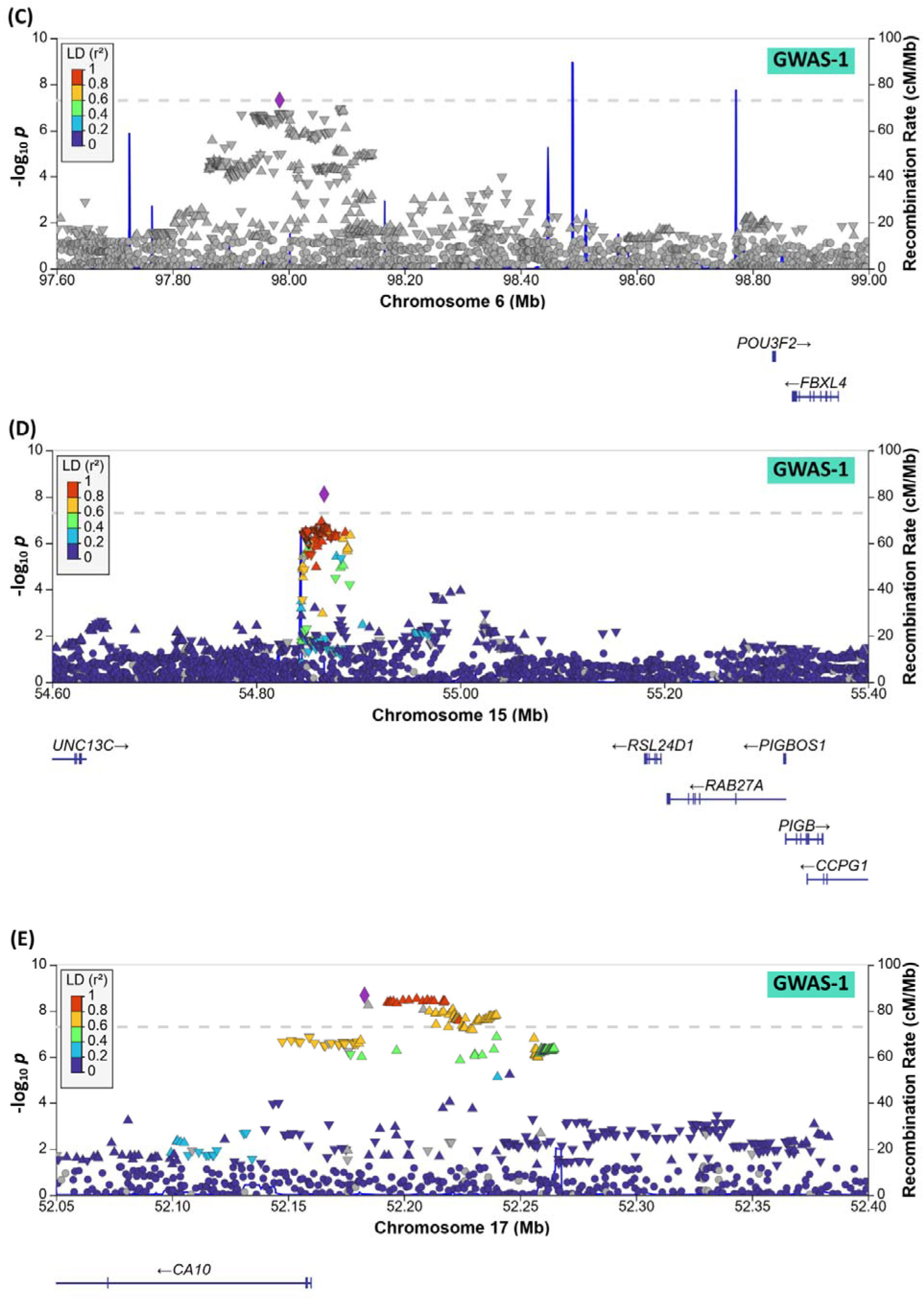
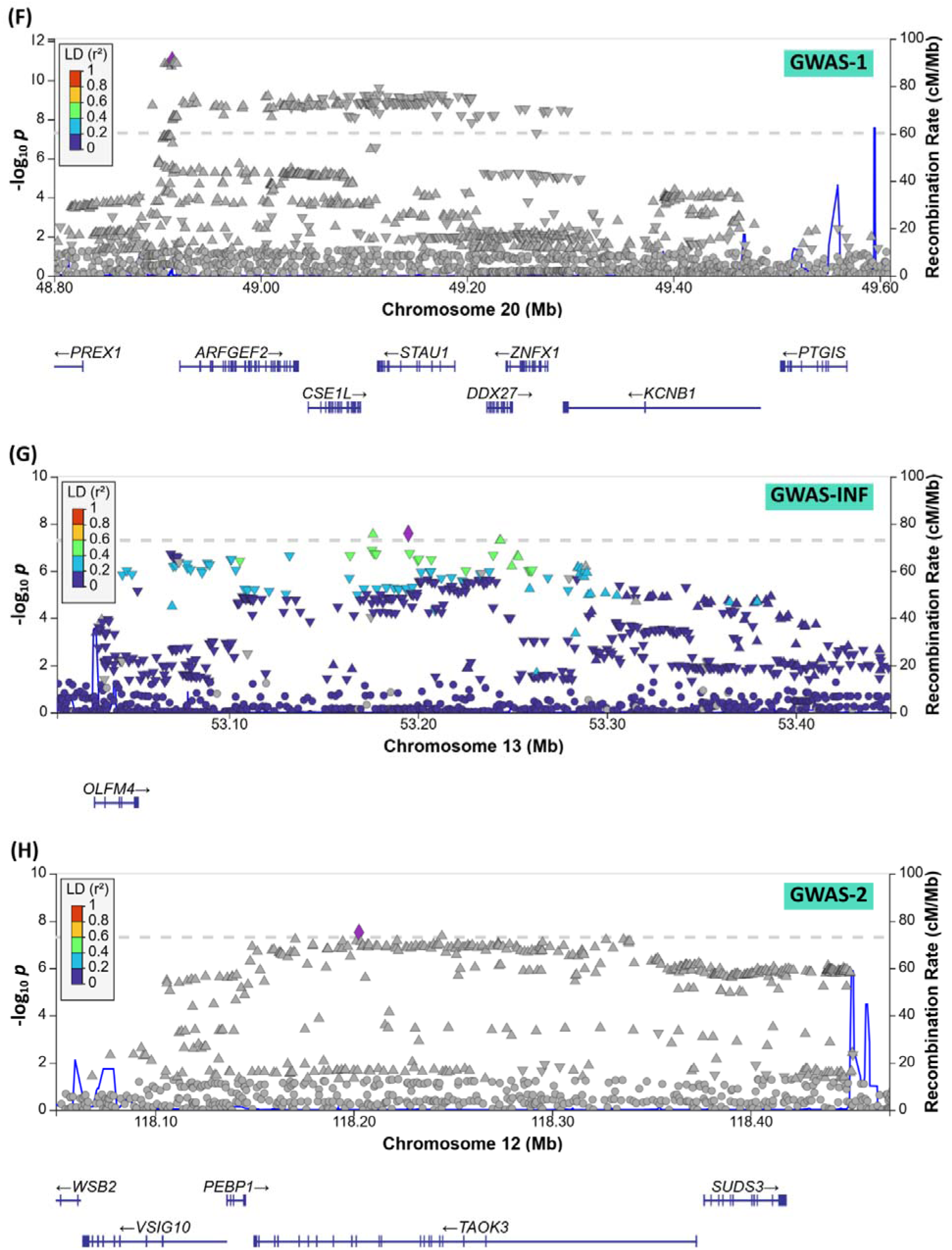
LocusZoom representations of the eight ME/CFS significantly associated loci in GWAS-1, GWAS-Infection and GWAS-2 (A-H). Significance of association (-log_10_*p*) is indicated on the left-hand *y*-axis, and chromosomal location on the *x*-axis. DNA variants are coloured by their linkage disequilibrium with the lead variant (purple diamond). The horizontal dashed lines indicate a genome-wide significance threshold of *p* < 5×10^−8^. Exon-intron representations of protein-coding genes are shown below. Recombination rates are indicated in blue (right-hand *y*-axis).

**Fig. S6:**
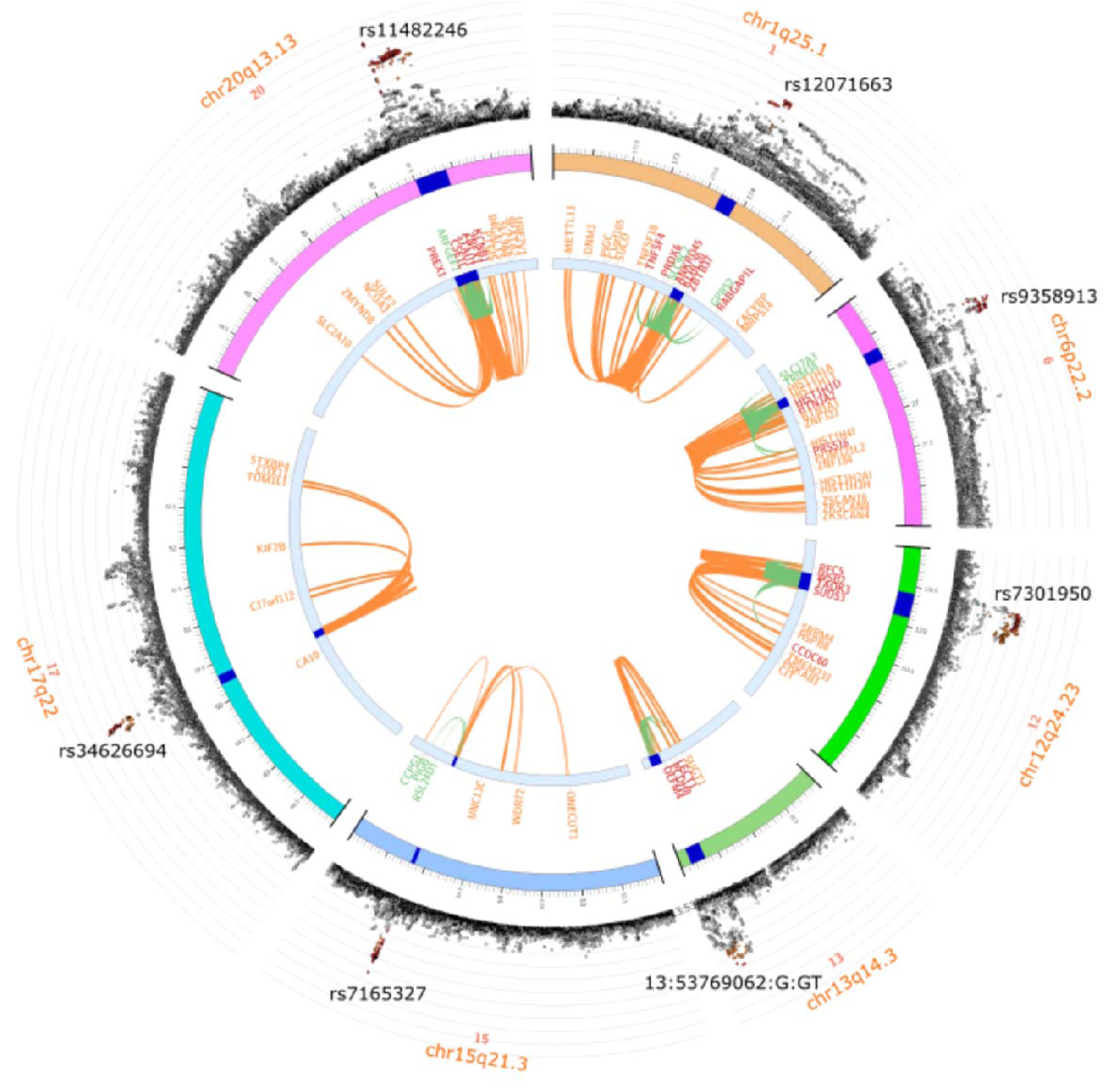
Candidate genes mapped to seven ME/CFS-associated chromosomal intervals by eQTL or chromatin interactions evidence. The outermost layer shows a Manhattan plot of variants within or near seven ME/CFS-associated chromosomal intervals; the eighth (chr6q16.1) has no reported expression quantitative trait loci (eQTLs) or chromatin interactions. Radial distance represents the-log_10_*p* of variants. Variants within genomic risk loci are colour-coded by linkage disequilibrium to independent significant variants: red (R^2^ > 0.8), orange (R^2^ > 0.6), green (R^2^ > 0.4) and blue (R^2^ > 0.2); others are shown in grey. The rsIDs of the lead variants are displayed within the outermost layer. The next layer in shows chromosomal location. Genomic risk loci are highlighted in blue. The innermost circle indicates genes only mapped by chromatin interactions (orange) or only by eQTLs (green). When a gene is mapped by both, its symbol is shown in red.

**Fig S7:**
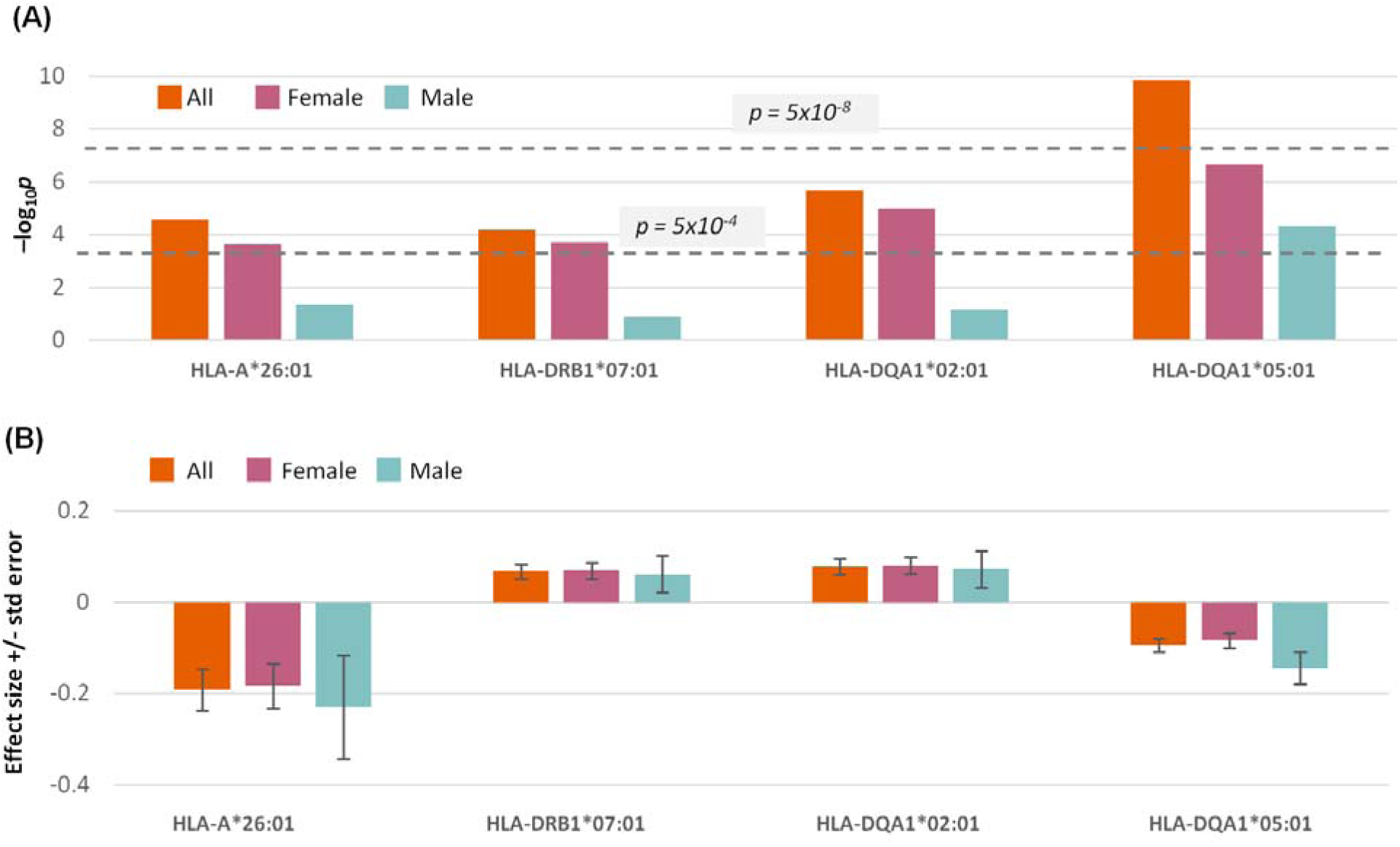
Association to ME/CFS of four human leukocyte antigen (HLA) alleles for all cases (shown in orange), females only (burgundy) or males only (green): one class I allele (HLA-A*26:01), and three class II alleles (HLA-DRB1*07:01, HLA-DQA1*02:01 and HLA-DQA*05:01). (A) Significance of association: –log_10_*p* (*y*-axis). Two *p*-value thresholds are shown as horizontal dashed lines: the genome-wide significance threshold, *p* = 5 x 10 (upper), and the Bonferroni-correction threshold, 5 x 10 (lower). (B) Association effect-sizes and their standard errors.

**Fig S8:**
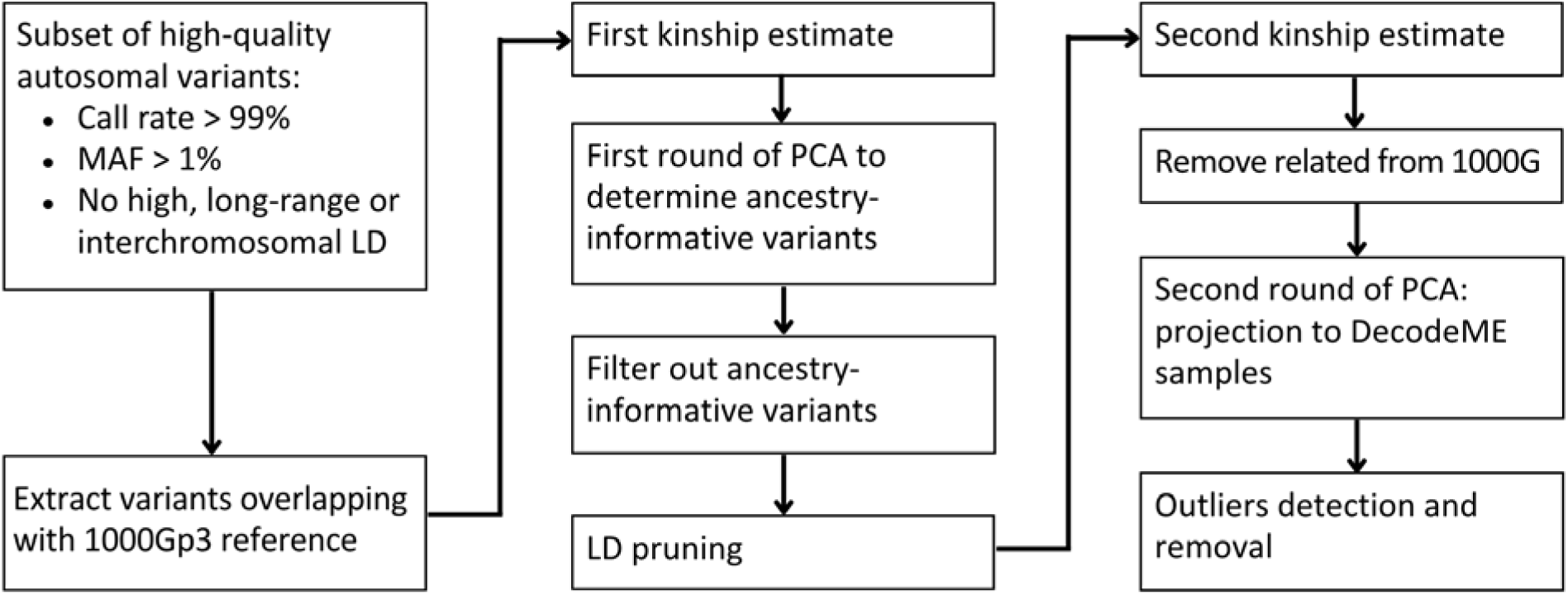
Flowchart of PCA-based ancestry grouping.

## CREDIT

Below we list contributor roles for our paper, using the National Information Standards Organization (NISO) Contributor Role Taxonomy (CRediT) (see https://credit.niso.org/).

| Name | Roles |
| --- | --- |
| Erik Abner | Resources |
| Øyvind Almelid | Resources, Software |
| Sumy V. Baby | Data Curation, Methodology, Resources |
| Tom Baker | Investigation, Data Curation, Formal Analysis, Resources, Software, Writing – Review & Editing |
| Helen Baxter | Investigation, Project Administration |
| Sjoerd Beentjes | Formal Analysis |
| Cindy Boer | Resources |
| Danielle Boobyer | Project Administration |
| Thibaud Boutin | Investigation, Methodology, Data Curation, Formal Analysis, Supervision, Visualization, Writing – Original Draft Preparation, Writing – Review & Editing |
| Andrew D. Bretherick | Methodology, Writing – Review & Editing |
| Sonya Chowdhury | Conceptualization, Funding Acquisition, Supervision, Writing – Review & Editing |
| Malgorzata Clyde | Investigation, Data Curation, Project Administration, Supervision, Writing – Review & Editing |
| Anne Connolly | Data Curation, Investigation, Project Administration, Writing – Original Draft Preparation, Writing – Review & Editing |
| Andy Devereux-Cooke | Conceptualization, Funding Acquisition, Supervision, Writing – Review & Editing |
| Joshua J. Dibble | Methodology, Formal Analysis, Visualization, Writing – Original Draft Preparation, Writing – Review & Editing |
| Claire Dransfield | Project Administration |
| Estonian Biobank Research Team | Resources |
| Esther Ewaoluwagbemiga | Formal Analysis |
| Diana Garcia | Investigation, Data Curation, Project Administration, Supervision, Writing – Review & Editing |
| Hele Haapaniemi | Resources |
| Gemma Hoyes | Writing – Review & Editing |
| John Ireland | Data Curation, Project Administration, Resources, Software |
| Shona M. Kerr | Conceptualization, Funding Acquisition, Methodology |
| Ava Khamseh | Formal Analysis |
| Daphne Lamirel | Writing – Review & Editing |
| Sian Leary | Conceptualization, Writing – Review & Editing |
| Isabel Lewis | Investigation, Project Administration |
| Ewan McDowall | Data Curation, Resources, Software |
| Simon J. McGrath | Conceptualization, Writing – Review & Editing |
| Julie Milton | Writing – Review & Editing |
| Nina Muirhead | Supervision |
| Emma Northwood | Conceptualization, Writing – Review & Editing |
| Hanna M. Ollila | Formal Analysis, Resources |
| David Perry | Data Curation, Resources, Software |
| Beth Pollack | Resources |
| Chris P. Ponting | Conceptualization, Formal Analysis, Funding Acquisition, Investigation, Methodology, Supervision, Visualization, Writing – Original Draft Preparation, Writing – Review & Editing |
| Ella Ponting | Project Administration, Writing – Review & Editing |
| Anna Redshaw | Conceptualization |
| Judith Rosmalen | Resources |
| Sirine Saafi | Resources |
| Gemma L. Samms | Formal Analysis, Methodology, Writing – Review & Editing |
| Richa Saxena | Resources |
| Charles Shepherd | Conceptualization |
| Nasa Sinnott-Armstrong | Resources |
| Joshua Slaughter | Formal Analysis |
| Anniina Tervi | Resources |
| Claire Tripp | Conceptualization, Writing – Review & Editing |
| Alice Turner | Project administration, Writing – Review & Editing |
| Lea Urpa | Resources |
| Jesse Valliere | Resources |
| David van Heel | Resources |
| Veronique Vitart | Conceptualization, Funding Acquisition, Methodology, Supervision, Writing – Review & Editing |
| Jim M. Wilson | Conceptualization |
| Jareth C. Wolfe | Data Curation, Resources, Software |

## Definition of roles

**Conceptualization** – Ideas; formulation or evolution of overarching research goals and aims.

**Data curation** – Management activities to annotate (produce metadata), scrub data and maintain research data (including software code, where it is necessary for interpreting the data itself) for initial use and later re-use.

**Formal analysis** – Application of statistical, mathematical, computational, or other formal techniques to analyze or synthesize study data.

**Funding acquisition** – Acquisition of the financial support for the project leading to this publication.

**Investigation** – Conducting a research and investigation process, specifically performing the experiments, or data/evidence collection.

**Methodology** – Development or design of methodology; creation of models.

**Project administration** – Management and coordination responsibility for the research activity planning and execution.

**Resources** – Provision of study materials, reagents, materials, patients, laboratory samples, animals, instrumentation, computing resources, or other analysis tools. Software – Programming, software development; designing computer programs; implementation of the computer code and supporting algorithms; testing of existing code components.

**Supervision** – Oversight and leadership responsibility for the research activity planning and execution, including mentorship external to the core team. Validation – Verification, whether as a part of the activity or separate, of the overall replication/reproducibility of results/experiments and other research outputs.

**Visualization** – Preparation, creation and/or presentation of the published work, specifically visualization/data presentation.

**Writing** – original draft – Preparation, creation and/or presentation of the published work, specifically writing the initial draft (including substantive translation).

**Writing** – review & editing – Preparation, creation and/or presentation of the published work by those from the original research group, specifically critical review, commentary or revision – including pre-or post-publication stages

## REFERENCES

1. Hanson MR, Germain A. Letter to the editor of metabolites. Vol. 10, Metabolites. MDPI AG; 2020.

2. Bretherick AD, McGrath SJ, Devereux-Cooke A, Leary S, Northwood E, Redshaw A, et al. Typing myalgic encephalomyelitis by infection at onset: A DecodeME study. NIHR open research. 2023;3:20.

3. Samms GL, Ponting CP. Unequal access to diagnosis of myalgic encephalomyelitis in England. BMC Public Health. 2025 Apr 22;25(1):1417.

4. Angelsen A, Schei T. EMEA survey of ME/CFS patients in Europe: Same disease, different approaches and experiences [Internet]. 2024 [cited 2024 Oct 29]. Available from: https://www.europeanmealliance.org/documents/emeaeusurvey/EMEAMEsurveyreport2024.pdf

5. Committee on the Diagnostic Criteria for Myalgic Encephalomyelitis/Chronic Fatigue Syndrome; Board on the Health of Select Populations; Institute of Medicine. Beyond Myalgic Encephalomyelitis/Chronic Fatigue Syndrome: Redefining an Illness. 2015.

6. Hickie I, Davenport T, Wakefield D, Vollmer-Conna U, Cameron B, Vernon SD, et al. Post-infective and chronic fatigue syndromes precipitated by viral and non-viral pathogens: Prospective cohort study. Br Med J. 2006 Sep 16;333(7568):575–8.

7. Katz BZ, Shiraishi Y, Mears CJ, Binns HJ, Taylor R. Chronic fatigue syndrome after infectious mononucleosis in adolescents. Pediatrics. 2009 Jul;124(1):189–93.

8. Komaroff AL, Lipkin WI. ME/CFS and Long COVID share similar symptoms and biological abnormalities: road map to the literature. Vol. 10, Frontiers in Medicine. Frontiers Media SA; 2023.

9. Cairns R, Hotopf M. A systematic review describing the prognosis of chronic fatigue syndrome. Occup Med (Lond). 2005 Jan;55(1):20–31.

10. Kielland A, Liu J. What can wage development before and after a G93.3 diagnosis tell us about prognoses for myalgic encephalomyelitis? Social Sciences & Humanities Open. 2025;11:101206.

11. Tyson S, Stanley K, Gronlund TA, Leary S, Emmans Dean M, Dransfield C, et al. Research priorities for myalgic encephalomyelitis/chronic fatigue syndrome (ME/CFS): the results of a James Lind alliance priority setting exercise. Fatigue. 2022;10(4):200–11.

12. Albright F, Light K, Light A, Bateman L, Cannon-Albright LA. Evidence for a heritable predisposition to chronic fatigue syndrome. BMC Neurol. 2011 May 27;11.

13. Underhill R, O’Gorman R. Prevalence of Chronic fatigue syndrome within families of CFS patients. J Chronic Fatige Syndr [Internet]. 2006 [cited 2024 Oct 29];13(3–13). Available from: https://www.tandfonline.com/doi/epdf/10.1300/J092v13n01_02?needAccess=true

14. Wang Q, Dhindsa RS, Carss K, Harper AR, Nag A, Tachmazidou I, et al. Rare variant contribution to human disease in 281,104 UK Biobank exomes. Nature. 2021 Sep 23;597(7877):527–32.

15. Spencer CCA, Su Z, Donnelly P, Marchini J. Designing genome-wide association studies: Sample size, power, imputation, and the choice of genotyping chip. PLoS Genet. 2009 May;5(5).

16. Hajdarevic R, Lande A, Mehlsen J, Rydland A, Sosa DD, Strand EB, et al. Genetic association study in myalgic encephalomyelitis/chronic fatigue syndrome (ME/CFS) identifies several potential risk loci. Brain Behav Immun. 2022 May 1;102:362–9.

17. Kurki MI, Karjalainen J, Palta P, Sipilä TP, Kristiansson K, Donner KM, et al. FinnGen provides genetic insights from a well-phenotyped isolated population. Nature [Internet]. 2023;613(7944):508–18. Available from: https://www.nature.com/articles/s41586-022-05473-8

18. Neale. Neale Lab UK Biobank [Internet]. [cited 2024 Nov 19]. Available from: https://www.nealelab.is/uk-biobank/

19. Canela-Xandri O, Rawlik K, Tenesa A. An atlas of genetic associations in UK Biobank. Nat Genet [Internet]. 2018;50(11):1593–9. Available from: https://www.nature.com/articles/s41588-018-0248-z

20. Zhou W, Nielsen JB, Fritsche LG, Dey R, Gabrielsen ME, Wolford BN, et al. Efficiently controlling for case-control imbalance and sample relatedness in large-scale genetic association studies. Nat Genet. 2018 Sep 1;50(9):1335–41.

21. Dibble JJ, McGrath SJ, Ponting CP. Genetic risk factors of ME/CFS: A critical review. Vol. 29, Human Molecular Genetics. Oxford University Press; 2020. p. R118–25.

22. Fry A, Littlejohns TJ, Sudlow C, Doherty N, Adamska L, Sprosen T, et al. Comparison of Sociodemographic and Health-Related Characteristics of UK Biobank Participants with Those of the General Population. Am J Epidemiol. 2017 Nov 1;186(9):1026–34.

23. Liles EG, Irving SA, Koppolu P, Crane B, Naleway AL, Brooks NB, et al. Classification Accuracy and Description of Myalgic Encephalomyelitis/Chronic Fatigue Syndrome in an Integrated Health Care System, 2006–2017. Perm J. 2024 Jun 19;1–12.

24. Devereux-Cooke A, Leary S, McGrath SJ, Northwood E, Redshaw A, Shepherd C, et al. DecodeME: community recruitment for a large genetics study of myalgic encephalomyelitis / chronic fatigue syndrome. BMC Neurol. 2022 Dec 1;22(1).

25. Bycroft C, Freeman C, Petkova D, Band G, Elliott LT, Sharp K, et al. The UK Biobank resource with deep phenotyping and genomic data. Nature. 2018 Oct 11;562(7726):203–9.

26. Carruthers BM, Jain AK, De Meirleir KL, Peterson DL, Klimas NG, Lerner AM, et al. Myalgic Encephalomyelitis/Chronic Fatigue Syndrome. J Chronic Fatigue Syndr. 2003 Jan 4;11(1):7–115.

27. Mbatchou J, Barnard L, Backman J, Marcketta A, Kosmicki JA, Ziyatdinov A, et al. Computationally efficient whole-genome regression for quantitative and binary traits. Nat Genet. 2021 Jul 1;53(7):1097–103.

28. Price AL, Patterson NJ, Plenge RM, Weinblatt ME, Shadick NA, Reich D. Principal components analysis corrects for stratification in genome-wide association studies. Nat Genet. 2006 Aug;38(8):904–9.

29. Privé F, Luu K, Blum MGB, McGrath JJ, Vilhjálmsson BJ. Efficient toolkit implementing best practices for principal component analysis of population genetic data. Bioinformatics. 2020 Aug 15;36(16):4449–57.

30. Watanabe K, Taskesen E, Van Bochoven A, Posthuma D. Functional mapping and annotation of genetic associations with FUMA. Nat Commun. 2017 Dec 1;8(1).

31. Wallace C, Giambartolomei C, Plagnol V. coloc: Colocalisation Tests of Two Genetic Traits. https://cran.r-project.org/web/packages/coloc/index.html. 2023.

32. Giambartolomei C, Vukcevic D, Schadt EE, Franke L, Hingorani AD, Wallace C, et al. Bayesian test for colocalisation between pairs of genetic association studies using summary statistics. PLoS Genet. 2014 May;10(5):e1004383.

33. Bulik-Sullivan B, Loh PR, Finucane HK, Ripke S, Yang J, Patterson N, et al. LD score regression distinguishes confounding from polygenicity in genome-wide association studies. Nat Genet. 2015 Feb 25;47(3):291–5.

34. Sijtsma A, Rienks J, van der Harst P, Navis G, Rosmalen JGM, Dotinga A. Cohort Profile Update: Lifelines, a three-generation cohort study and biobank. Int J Epidemiol. 2022 Oct 13;51(5):e295–302.

35. Samms GL, Ponting CP. Defining a High-Quality Myalgic Encephalomyelitis/Chronic Fatigue Syndrome cohort in UK Biobank. NIHR open research. 2025;5:39.

36. Ofcom. Online Nation 2024 report [Internet]. 2024 [cited 2025 Apr 22]. Available from: https://www.ofcom.org.uk/siteassets/resources/documents/research-and-data/online-research/online-nation/2024/online-nation-2024-report.pdf?v=386238

37. Jones CL, Younger J. Possible Racial Disparities in the Diagnosis of Myalgic Encephalomyelitis/Chronic Fatigue Syndrome (ME/CFS). Int J Environ Res Public Health. 2025 Feb 14;22(2).

38. Pardhan S, Sehmbi T, Wijewickrama R, Onumajuru H, Piyasena MP. Barriers and facilitators for engaging underrepresented ethnic minority populations in healthcare research: an umbrella review. Int J Equity Health. 2025 Mar 12;24(1):70.

39. NICE. Myalgic encephalomyelitis (or encephalopathy)/chronic fatigue syndrome: diagnosis and management NICE guideline [Internet]. 2021. Available from: www.nice.org.uk/guidance/ng206

40. Randall JC et al. Sex-stratified genome-wide association studies including 270,000 individuals show sexual dimorphism in genetic loci for anthropometric traits. 2013 [cited 2025 Jun 26]; Available from: https://pubmed.ncbi.nlm.nih.gov/23754948/

41. Harlow CE, Uzochukwu E, Fernando HA, Mordaunt CE, Hughey JM, Eicher JD, et al. GWAS of Extended Prescription Analgesic Use Identifies Novel Genetic Loci in Chronic Pain [Internet]. 2024 [cited 2025 Jul 24]. Available from: https://www.medrxiv.org/content/10.1101/2024.12.02.24318312v1

42. de Leeuw CA, Mooij JM, Heskes T, Posthuma D. MAGMA: Generalized Gene-Set Analysis of GWAS Data. PLoS Comput Biol. 2015 Apr 1;11(4).

43. Qi T, Song L, Guo Y, Chen C, Yang J. From genetic associations to genes: methods, applications, and challenges. Trends Genet. 2024 Aug;40(8):642–67.

44. Marzluff et al. The Human and Mouse Replication-Dependent Histone Genes. 2002 [cited 2025 Jun 26]; Available from: https://pubmed.ncbi.nlm.nih.gov/12408966/

45. Minowa-Nozawa A, Nozawa T, Murase K, Nakagawa I. RabGAP1L modulates Rab7A and Rab10 to orchestrate cell-autonomous immunity. Cell Rep. 2025 May 27;44(5):115599.

46. Fernbach S, Spieler EE, Busnadiego I, Karakus U, Lkharrazi A, Stertz S, et al. Restriction factor screening identifies RABGAP1L-mediated disruption of endocytosis as a host antiviral defense. Cell Rep. 2022 Mar 22;38(12):110549.

47. Mohammed F, Willcox CR, Willcox BE. A Brief Molecular History of Vγ9Vδ2 TCR-Mediated Phosphoantigen Sensing. Vol. 331, Immunological Reviews. John Wiley and Sons Inc; 2025.

48. Sarter K, Leimgruber E, Gobet F, Agrawal V, Dunand-Sauthier I, Barras E, et al. Btn2a2, a T cell immunomodulatory molecule coregulated with MHC class II genes. J Exp Med. 2016 Feb 8;213(2):177–87.

49. Frech M, Danzer H, Uchil P, Azizov V, Schmid E, Schälter F, et al. Butyrophilin 2a2 (Btn2a2) expression on thymic epithelial cells promotes central T cell tolerance and prevents autoimmune disease. J Autoimmun. 2023 Sep;139:103071.

50. Hu MM, Yang Q, Xie XQ, Liao CY, Lin H, Liu TT, et al. Sumoylation Promotes the Stability of the DNA Sensor cGAS and the Adaptor STING to Regulate the Kinetics of Response to DNA Virus. Immunity. 2016 Sep 20;45(3):555–69.

51. Shen J, Lai W, Li Z, Zhu W, Bai X, Yang Z, et al. SDS3 regulates microglial inflammation by modulating the expression of the upstream kinase ASK1 in the p38 MAPK signaling pathway. Inflamm Res. 2024 Sep;73(9):1547–64.

52. Montoliu-Gaya L, Tietze D, Kaminski D, Mirgorodskaya E, Tietze AA, Sterky FH. CA10 regulates neurexin heparan sulfate addition via a direct binding in the secretory pathway. EMBO Rep. 2021 Apr 7;22(4):e51349.

53. Zhang P, Lu H, Peixoto RT, Pines MK, Ge Y, Oku S, et al. Heparan Sulfate Organizes Neuronal Synapses through Neurexin Partnerships. Cell. 2018 Sep 6;174(6):1450–1464.e23.

54. Lin TB, Lai CY, Hsieh MC, Jiang JL, Cheng JK, Chau YP, et al. Neuropathic Allodynia Involves Spinal Neurexin-1β-dependent Neuroligin-1/Postsynaptic Density-95/NR2B Cascade in Rats. Anesthesiology. 2015 Oct;123(4):909–26.

55. Kutay U, Bischoff FR, Kostka S, Kraft R, Görlich D. Export of importin alpha from the nucleus is mediated by a specific nuclear transport factor. Cell. 1997 Sep 19;90(6):1061–71.

56. Gao CL, Song JQ, Yang ZN, Wang H, Wu XY, Shao C, et al. Chemoproteomics of Marine Natural Product Naamidine J Unveils CSE1L as a Therapeutic Target in Acute Lung Injury. J Am Chem Soc. 2024 Oct 16;146(41):28384–97.

57. Islam A, Shen X, Hiroi T, Moss J, Vaughan M, Levine SJ. The brefeldin A-inhibited guanine nucleotide-exchange protein, BIG2, regulates the constitutive release of TNFR1 exosome-like vesicles. Journal of Biological Chemistry. 2007 Mar 30;282(13):9591–9.

58. Wang Y, Yuan S, Jia X, Ge Y, Ling T, Nie M, et al. Mitochondria-localised ZNFX1 functions as a dsRNA sensor to initiate antiviral responses through MAVS. Nat Cell Biol. 2019 Nov;21(11):1346–56.

59. Soskic B, Cano-Gamez E, Smyth DJ, Ambridge K, Ke Z, Matte JC, et al. Immune disease risk variants regulate gene expression dynamics during CD4+ T cell activation. Nat Genet. 2022 Jun;54(6):817–26.

60. Bonnen PE, Yarham JW, Besse A, Wu P, Faqeih EA, Al-Asmari AM, et al. Mutations in FBXL4 cause mitochondrial encephalopathy and a disorder of mitochondrial DNA maintenance. Am J Hum Genet. 2013 Sep 5;93(3):471–81.

61. Vandenberghe-Dürr S, Gilliet M, Di Domizio J. OLFM4 regulates the antimicrobial and DNA binding activity of neutrophil cationic proteins. Cell Rep. 2024 Oct 22;43(10):114863.

62. Smith MD, Harley ME, Kemp AJ, Wills J, Lee M, Arends M, et al. CCPG1 Is a Non-canonical Autophagy Cargo Receptor Essential for ER-Phagy and Pancreatic ER Proteostasis. Dev Cell. 2018 Jan 22;44(2):217–232.e11.

63. Li J, Gao E, Xu C, Wang H, Wei Y. ER-Phagy and Microbial Infection. Front Cell Dev Biol. 2021;9:771353.

64. Wray NR, Ripke S, Mattheisen M, Trzaskowski M, Byrne EM, Abdellaoui A, et al. Genome-wide association analyses identify 44 risk variants and refine the genetic architecture of major depression. Nat Genet. 2018 May;50(5):668–81.

65. Okbay A, Baselmans BML, De Neve JE, Turley P, Nivard MG, Fontana MA, et al. Genetic variants associated with subjective well-being, depressive symptoms, and neuroticism identified through genome-wide analyses. Nat Genet. 2016 Jun;48(6):624–33.

66. Hu J, Lupton MK, Byrne EM, Martin NG, Whiteman DC, Olsen CM, et al. Unravelling Sex Differences in the Genetic Architecture of Anxiety: A Genome-Wide Association Study in the UK Biobank [Internet]. 2025 [cited 2025 Jul 24]. Available from: https://www.medrxiv.org/content/10.1101/2025.02.09.25321968v1

67. Friligkou E, Løkhammer S, Cabrera-Mendoza B, Shen J, He J, Deiana G, et al. Gene discovery and biological insights into anxiety disorders from a large-scale multi-ancestry genome-wide association study. Nat Genet. 2024 Oct;56(10):2036–45.

68. Minikel EV, Painter JL, Dong CC, Nelson MR. Refining the impact of genetic evidence on clinical success. Nature. 2024 May 16;629(8012):624–9.

69. Abdellaoui A, Yengo L, Verweij KJH, Visscher PM. 15 years of GWAS discovery: Realizing the promise. Vol. 110, American Journal of Human Genetics. Cell Press; 2023. p. 179–94.

70. Nacul LC, Lacerda EM, Pheby D, Campion P, Molokhia M, Fayyaz S, et al. Prevalence of myalgic encephalomyelitis/chronic fatigue syndrome (ME/CFS) in three regions of England: a repeated cross-sectional study in primary care. BMC Med. 2011 Jul 28;9:91.

71. Griffith JP, Zarrouf FA. A systematic review of chronic fatigue syndrome: don’t assume it’s depression. Prim Care Companion J Clin Psychiatry. 2008;10(2):120–8.

72. Lammi V, Nakanishi T, Jones SE, Andrews SJ, Karjalainen J, Cortés B, et al. Genome-wide association study of long COVID. Nat Genet. 2025 Jun;57(6):1402–17.

73. Decode ME Researcher Access [Internet]. 2024 [cited 2025 Jul 24]. Available from: https://www.decodeme.org.uk/researcher-access/

74. Froehlich L, Hattesohl DB, Cotler J, Jason LA, Scheibenbogen C, Behrends U. Causal attributions and perceived stigma for myalgic encephalomyelitis/chronic fatigue syndrome. J Health Psychol. 2022 Sep;27(10):2291–304.

75. Kurki MI, Karjalainen J, Palta P, Sipilä TP, Kristiansson K, Donner KM, et al. FinnGen provides genetic insights from a well-phenotyped isolated population. Nature. 2023 Jan;613(7944):508–18.

