## Supplementary Methods for "Initial findings from the DecodeME genome-wide association study of myalgic encephalomyelitis/chronic fatigue syndrome"

### Contents


Co-production ... 2

Recruitment ... 2

Questionnaires ... 4

Inclusion criteria ... 5

DNA sample collection ... 6

Genotyping ... 7

Quality control steps and further analysis ... 7

Genotype calling ... 7

Merged case-control array genotypes ... 11

Phasing and genotype imputation ... 11

HLA imputation ... 12

Association testing ... 13

Replication analyses ... 15

Post-GWAS analyses ... 16

Table S10. Primary care codes. ... 17

Table S12: DecodeME Participant Numbers across the Workflow ... 19

REFERENCES ... 20

### Co-production

DecodeME was co-produced with patients, carers, and charities from the start. The project arose from the CFS/ME Research Collaborative (CMRC), which had already included patients and carers as full members of its executive board and demonstrated their value in 2018 (1), when preliminary discussions were being held with DecodeME's eventual funders.

ME/CFS has no effective treatment, and so people with it often have severely disabling symptoms that make taking part in a genetics study difficult. These include profound exhaustion, post-exertional malaise (being much more ill after physical or mental activity), 'brain fog', orthostatic intolerance (the inability to maintain an upright posture), and intolerance to light and sound (2). This meant that having the insights of people with ME/CFS and their carers to inform all aspects of our study was vital to make the study possible and to ensure that the views of people with the illness were represented.

At every stage of the study, therefore, people with lived experience jointly delivered DecodeME alongside professional researchers, including writing the grant proposal, co-authoring this and other DecodeME publications, taking part on an equal footing with academic team members in weekly, monthly and quarterly meetings, and contributing two of the three members of the project's decision-making group.

DecodeME's patient and public involvement (PPI) followed UK National Standards (3) and rooted our decisions in the priorities of people with lived experience of ME/CFS. PPI benefited the study's planning, design, and delivery in many ways, including by boosting recruitment and making the study's questionnaire easier for participants to understand (4). The project's approach to PPI has been held up as an example of good practice by UK Research and Innovation (5).

We intend to discuss DecodeME's co-production design and impact in detail in a future publication.

### Recruitment

#### Recruitment target

DecodeME was awarded funding in June 2020, by which time the COVID-19 pandemic was under way. We had originally aimed to collect DNA from at least 20,000 participants, because we estimated that this would give us 80% power to detect significant ($p < 5 \times 10^{-8}$) associations for common (minor allele frequency ≥ 5%) variants with modest (~1.1) relative risk. In response to the pandemic, we aimed to include 5,000 more participants who would be diagnosed with ME/CFS following SARS-CoV-2 virus infection.

We expected to lose participants at each stage (due to them not completing questionnaires, not meeting the DNA selection criteria, or not returning sample collection kits), and we estimated that to reach our sample target, we would need about 50,000 potential participants to initially engage.

### Recruitment methods

We decided early that direct recruitment of ME/CFS cases would be more successful and would make DecodeME more accessible to people with ME/CFS, compared with the alternative of indirect recruitment through the few specialised ME/CFS clinics, for several reasons. People with ME/CFS often report dismissive attitudes, stigma, and misconceptions from health professionals (6), and so might be less willing to sign up to a study recruiting through a clinic. Many people with ME/CFS are housebound or bedbound and so cannot travel to the clinics (7). Even those able to travel to a clinic face exhaustion, possible sensory overload, and the risk of post-exertional malaise and a relapse, all of which are a strong motivation not to attend. Finally, we wanted to recruit across the whole of the UK and not only in the regional centres where many clinics are located.

In order to recruit as many people with ME/CFS as possible, we committed to maximising understanding, transparency and communication, to build trust; and aimed to minimise how much time and energy participants would need to engage, in order to make it as easy as possible for them to take part in the study.

We therefore chose a 'spit and post' design in which we would mail saliva DNA collection kits to participants so that they could take part from home. We also decided to make it easier to take part in the study by having internet- and non-internet-based options for communicating and for completing the study questionnaires. We advertised these adaptations widely in an attempt to boost recruitment.

When funding was awarded in June 2020, the news spread via traditional and social media. Within ten days, 16,532 UK residents with a self-reported diagnosis of ME/CFS had pre-registered via our website to take part in DecodeME. It took two more years to acquire ethics approval, arrange contracts with suppliers and service providers (4), build the participant data infrastructure, and carry out a trial of sample collection. We publicly launched recruitment of participants on 12 September 2022. We closed recruitment on 31 January 2024, which was the final deadline for the return of saliva collection kits.

We used nine marketing and communications methods for recruitment:

1. We created a website as a study sign-up and information portal in 2020, before the announcement of the study's funding. We chose the branding and colours to make the site as accessible as possible for people with ME/CFS who were intolerant to light, and we enabled the site to be viewed in dark mode.
2. We emailed the 28,490 people who had pre-registered online since funding was announced and invited them to take part in the study.
3. A press relations agency (Hot Tin Roof) and the University of Edinburgh communications team secured media interviews for members of the DecodeME team with national and local media coverage to announce that we were recruiting people diagnosed with ME/CFS.

4. We hired Itineris, a digital marketing company, to help develop and carry out a targeted online advertising campaign to reach and enrol ('convert') eligible people with ME/CFS.
5. We used social media channels extensively, including Facebook, X (Twitter), Instagram and TikTok. We recruited 306 people with ME/CFS as social media ambassadors who created engaging content, including video. We also worked with influential and well-networked people in the ME/CFS community to maximise the project's online reach.
6. We created packs of tailored promotional information and materials, which we shared with charities, local support groups and clinicians for them to share with their networks and via their newsletters and websites.
7. We held live Question and Answer webinars. People with ME/CFS may struggle to concentrate on webinars in real-time, or may run out of energy to watch them, or may find them difficult to watch and listen due to intolerance of light and/or sound. To make the webinars accessible, we uploaded recordings of them to DecodeME's website and YouTube channel, including as audio-only and as subtitled videos, all with full transcripts.
8. We gave regular updates to all registered interested parties via an emailed or posted newsletter, and as blogs on our website. We also used these channels to support follow-up campaigns, including the sharing of the preliminary results of the first questionnaire, the last call for sign-up, and the deadline for returning 'spit-kits'.
9. We sent up to three email reminders to participants who started the process but did not progress to giving consent or completing the questionnaire, to encourage them to complete the remaining steps. Throughout the recruitment and DNA sample collection phase, we supported participants via a detailed Frequently Asked Questions section on our website and gave both email and phone support to participants, and potential participants who had questions or needed further help. We also used social media to answer specific questions. We actively monitored and managed all of these channels. We sent over 17,000 emails during recruitment in response to questions on a wide range of topics.

### Questionnaires

#### (i) Questionnaire content

We sent all potential study participants a questionnaire that included symptom questions to allow us to apply criteria based on the Canadian Consensus criteria (CCC) and Institute of Medicine (IOM) / National Academy of Medicine (NAM) criteria for ME/CFS (see below) (2,8). There were also questions about sex assigned at birth, ethnic group, and blood group (if known), to help with genotyping quality-control.

We trialled a draft of the questionnaire, developed with PPI, on 50 volunteers. Of 41 respondents, 16 (39%) reported problems, leading to them not answering some questions, selecting too many answers, or adding clarifying comments. Having clear, easily understood questions was important, given that people with ME/CFS have limited energy and may have cognitive processing problems due to their illness, and so we redrafted the questionnaire accordingly.

We trialled the online version on 470 randomly sampled pre-registrants and 88 (19%) reported problems with it. Their feedback revealed minor issues, which we addressed.

We invited participants to complete an optional second questionnaire between December 2023 and February 2024. This enabled us to collect detailed phenotype data using questions that we had not included in the first questionnaire in order to keep the burden on participants as low as possible. We plan to report responses to this second questionnaire in a separate paper. Our DecodeME questionnaires are available under a CC-BY license from the Open Science Framework, at https://osf.io/rgqs3/files/osfstorage.

**(ii) Adaptations and support for completing questionnaires**

Participants could complete questionnaires with the help of family or friends (9). We offered the questionnaire in both online and paper formats, because some people with ME/CFS are intolerant to light and struggle to use screens. They could also get support via telephone from Helen Baxter of the 25% ME Group, a UK charity that provides services to severely ill people with ME/CFS (9).

We designed the online questionnaire to save responses if the participant needed to complete it in short sessions with breaks between, rather than all at once. Each web page showed only a limited number of questions to avoid being overwhelming.

Participants could also request a paper copy of the questionnaire to complete at home, again with the help of family or carers if needed. We posted these paper questionnaires to participants, who could return them postage-paid.

**(iii) Questionnaire data storage**

We collected questionnaire data via the online Qualtrics™ platform and downloaded it into a relational database stored in a secure location at the University of Edinburgh.

### Inclusion criteria

To be included in DecodeME, participants had to live in the UK and be at least 16 years old. To be a DecodeME 'case', who could give a DNA sample for the GWAS, participants also had to have a diagnosis of ME, CFS, ME/CFS or CFS/ME from a health professional. Further, they had to meet criteria that we based on the CCC and/or IOM/NAM diagnostic criteria (2,8):

**IOM/NAM-based criteria.** Participants had to **meet all four** of the following criteria:

- Have had symptoms for over six months, but not over their entire life.
- Have frequent fatigue that can worsen when active, that reduces their activities, and feels 'like a battery that can never fully recharge even when I rest'.
- Have reported unrefreshing sleep, and post-exertional malaise (new symptoms, or the worsening of symptoms, after physical or mental exertion that would not have caused a problem before the illness) that reduces how much they can do over a long time, which can be for over a day.
- Have reported cognitive problems and/or orthostatic intolerance.

**CCC-based criteria.** Participants had to **meet all six** of the following criteria:

- Have experienced fatigue often, which worsens when they're active and that reduces their activities, and which affects them both physically and mentally.
- Have reported post-exertional malaise (new symptoms, or the worsening of symptoms, after physical, mental, or emotional exertion that would not have caused a problem before the illness) and that reduces how much they can do over a long time, which can be for over a day.
- Have unrefreshing or problematic sleep.
- Have pain.
- Have two or more neurological or cognitive symptoms (e.g., 'brain fog').
- Have one or more symptoms from two sets of autonomic, neuroendocrine, or immune symptoms.

The CCC (2) exclude people as ME/CFS cases if they report any of 25 active medical conditions that could explain most of the major symptoms. We therefore initially excluded participants who reported that any of these conditions had given them symptoms in the preceding six months. However, the DecodeME Scientific Advisory Board later proposed removing these exclusionary criteria to help DecodeME meet its participant recruitment target and to avoid disproportionately excluding people of lower socioeconomic status, who tend to have worse overall health (10).

We agreed this proposal across DecodeME and implemented it from 26 June 2023, requesting saliva samples from over 7,800 participants whose saliva DNA we had not originally asked for. In case this meant that we had included participants who did not have ME/CFS, we added planned sensitivity analyses to test whether a statistically significant genetic signal that colocalises with ME/CFS arose only from subsets of cases who disproportionately reported a particular comorbidity. No significant colocalisation with a comorbid condition has yet been identified, so these sensitivity analyses have not been required.

### DNA sample collection

We designed DNA sample collection to be as straightforward and accessible as possible for people with ME/CFS (4). We used a fulfilment company (Gemini Print) to mail Isohelix GeneFiX™ saliva sample kits to eligible participants, so that they could contribute their 2ml sample at home and post it to the UK Biocentre for DNA extraction.

A fulfilment company prepared the kits. Each contained a collection tube and funnel, a sealable sample safety bag and absorbent pad, a shipping box and bag, and a prepaid postage label. We also included a simple instruction leaflet that we created, which explained how to provide and mail a useable sample, plus a link to an online video that showed how to use the kit. Sample packages met Royal Mail guidelines to be classed as 'exempt human specimens' so that participants or their helpers could post them in any post box.

We tested the kit supply and sample return processes before the recruitment launch.

### Genotyping

The UK Biocentre extracted DNA using the Kingfisher system, or, for 661 samples, the Promega Heater Shaker Magnet instrument. The extracted DNA and samples were quality-assessed and stored. If samples did not meet the required quality standard (producing less than 50 ng/µL of DNA in both of two extraction attempts), we emailed or wrote to the participant to explain that we needed a second sample and sent another sample kit.

The UK Biocentre quantified the extracted DNA samples. If there was at least 50ng/µl of DNA in a sample, it was normalised to 60ng/µl, and stored at -80$^{o}$C in a 54µl primary aliquot. Any remaining volume was stored in a secondary aliquot. The primary aliquots were then plated in four batches of 4,980, 5,450, 5,075, and 2,761 samples respectively. They were further quantified and then sent to Thermo Fisher, who genotyped them with the UK Biobank (UKB) Axiom™ array. Across all four batches, 693 samples failed genotyping. Array intensity data in CEL format was returned for secure storage on the University of Edinburgh's servers. The secondary aliquots are in storage for future whole genome sequencing.

### Quality control steps and further analysis

All data quality control (QC) and analysis followed a pre-registered Data Analysis Plan (11), later updated (12).

### Genotype calling

We called genotypes by batch using the Axiom Analysis suite (AxAS v5.4), following the *Best Practices Genotyping Analysis Workflow* option and inbuilt library, Axiom_UKB_WCSG.r5. On a first calling round, Thermo Fisher applied the recommended ('default') thresholds to quality check both samples and markers as described in (13). We ran the same genotype analysis workflow again, but without samples that failed QC, and changing variant filtering thresholds for Fisher's linear discriminant (FLD), Heterozygous Strength Offset and Homozygote Ratio Offset to ≥ 4.5, ≥ 0 and ≥ 0, respectively. Variants classified as 'off-target', 'CallRateBelowThreshold', 'Other' and 'Unexpected genotype frequency' were discarded. Genotype clusters were visually inspected, especially for sex-linked markers, autosomal variants with strong departure from Hardy-Weinberg equilibrium (HWE), and for rarer variants that are more likely to be less well genotyped or to present problematic clustering.

**Plate and batch effects**

We next accounted for possible plate and batch effects. We tested for differences in genotype frequencies between plates within a single batch (plate effect) for each remaining variant using a Fisher's exact test on the 2 × 3 contingency table of genotype counts following UKB methodology (14). With four batches, 197 plates and 836,727 markers, we applied a Bonferroni corrected *p*-value threshold of $7 \times 10^{-11}$ (corresponding to a family-wise error rate < 0.005) to indicate a significant plate effect. We filtered out variants that met this threshold.

We tested genotyping consistency across the four batches using case-case GWAS (PLINK 2.0, glm model), for unrelated European individuals' samples, once we had defined them, with data from each batch tested against data in all other batches. We visually inspected genotyping clusters for variants showing batch-association with $p < 10^{-5}$ and we removed poorly clustered variants (11) from downstream analysis.

**Concordance**

Each plate had two control wells with DNA samples from two individuals (NA19315, NA19318) from the African superpopulation (AFR) of the 1000 Genomes project (15). Samples from some cases were present on multiple plates. We tested such duplicate samples for concordance. If discordance (*d*), as defined by (14), was more than 0.05 for one or both of the control samples, then we flagged and excluded the marker. We also did this analysis for known DecodeME sample duplicates present in different batches and excluded variants that were discordant across batches.

**Identifying females and males**

We inferred the genetic sex of participants during the automated genotype calling process in AxAS based on chromosome X- and Y- linked variants. For samples that failed automated sex-inference, we visualised probeset intensities across sex chromosomes using the AxAS Copy Number Variant workflow and compared them to those of male or female references (16). We removed samples if they showed sex-chromosome aneuploidy, or if recorded sex and genetically inferred sex were different (in case this reflected a data error).

**Ancestry and kinship**

We selected a subset of high-quality variants for ancestry and kinship estimation by further removing: (i) non-autosomal variants; (ii) A/T or G/C DNA variants that can introduce DNA strand ambiguity when merging genotype data with the ancestry reference panel; (iii) samples with a call rate < 0.95; (iv) DNA variants with a call rate < 0.99; (v) DNA variants with a minor allele frequency (MAF) < 0.01; and, (vi) DNA variants in high linkage disequilibrium (LD), as listed in Bycroft et al. (14).

We identified and removed additional long-range LD using the *R* package *bigsnpr* (17). In brief, we estimated relatedness among participants using KING (18) with options '*--unrelated*' and '*--degree 3*' (that is, we considered pairs of samples to be related when their kinship coefficient was more than 0.04419).

We identified non-ancestry-informative variants (PC loadings < 0.03) after a first round of principal component analysis (PCA) using a reference population with representatives from all major ancestry groups (15), as described for UKB samples in (14).

We discarded samples identified as monozygotic twins or parent-child pairs whose reported years of birth were incompatible with such genetic relationships. We also discarded unexpected duplicate samples. Among technical duplicates from the same DecodeME participant, we kept the sample with the highest QC metric, using Dish QC (19), or if a tie, the QC call rate.

We inferred major group ancestries (African, American, East Asian, European, and South Asian) by PCA projection of DecodeME samples onto the 1000 Genome reference population samples (15) using *bigsnpr* (17) (**Fig. S8**). We completed a second round of PCA on the European ancestry cohort to calculate relatedness and principal components for downstream analyses. We ran the implementation of KING used in PLINK 2.0, on the subset of variants ('in_Relatedness') used by UKB that were present in DecodeME's data (68,453 SNPs). This was because not subsetting the SNPs used would have severely inflated relatedness estimates (14).

We identified four more participants with more than 200 predicted relations in the dataset. Following Bycroft *et al.* (14) we excluded them from downstream analyses after PCA calculation. No participants included in the list of 977 individuals who were excluded from kinship estimation by Bycroft et al. then remained in our dataset.

We called 72 UKB control samples as related in our analysis that were listed as unrelated by UKB. Nevertheless, their relatedness coefficients approached the threshold used (~0.0442), presumably due to slightly differing QC, and so we included them as related. This is a conservative approach, as these samples do not contribute to PC calculation and are only projected onto the PC axes. We again performed PCA using the method of Prive et al. (17)(20) which also accounts for regions of long-range LD when performing the singular value decomposition step. For this, we considered only the 72,192 SNPs used by UKB ('in_PCA') that were included in our data. As before, the PCA calculation removed the list of related individuals, calculated the PCs on the unrelated data, and then projected the related individuals onto these axes. We saw no outliers in our data and so removed no samples at this stage (21). In summary, PCA was performed on a dataset of 281,631 individuals, with 58,582 considered third-degree or closer relatives.

However, the resulting plots showed that some population stratification remained. By labelling UKB controls who had 'United Kingdom', 'Italy', or 'Poland' ancestries, we found that almost all outliers had unlabelled ancestries. This suggested a PC1 cutoff (PC1 > 23), which removed 7,093 UKB controls and 270 DecodeME cases. The remaining subset formed the GWAS-1 group (15,579 cases and 259,909 controls); we recalculated PCs, as described above, for this revised cohort.

**UK Biobank control samples**

We selected controls from the UKB as a general population cohort, but we excluded 5,365 UKB ME/CFS cases. These were defined as people who self-reported chronic fatigue syndrome ('CFS') in the baseline questionnaire (Data Field 20002), or who had electronic health record (EHR) evidence for ME/CFS (see below), or who self-reported CFS in the verbal interview at any of the four visits to the UKB assessment centre, or who answered 'Yes' to 'Ever had Chronic Fatigue Syndrome or Myalgic Encephalomyelitis (M.E.)' in the pain questionnaire (Data Field 120010). We excluded the 88 participants who self-reported CFS at baseline but answered 'No' to this question in the pain questionnaire. We also excluded participants who ambiguously answered 'Do not know' (N = 2,065) or 'Prefer not to answer' (N = 19).

We will exclude potential ME/CFS cases who reported both a CFS diagnosis and good health in the baseline questionnaire as either cases or controls for GWAS-3 or GWAS-4.

We used available EHR data (hospital in-patient data based on ICD-10 codes and GP primary care record information on referrals and diagnoses) to exclude more UKB individuals from acting as controls. The G93.3 ICD-10 code (Post-Viral Fatigue Syndrome) or R53 (Malaise and fatigue) are unlikely to be specific enough to identify people with ME/CFS as cases in the UKB. However, among the 1,393 UKB participants linked to the G93.3 code, most (63%, N = 878) self-reported having ME/CFS in the baseline or pain questionnaires. Similarly, over 50% (N = 801) of the 1,575 UKB participants linked to an ME/CFS-relevant primary care code (**Table S10**) self-reported ME/CFS in the UKB questionnaires. In comparison, only 0.10% (N = 508) of all UKB participants not self-reporting ME/CFS were linked to the G93.3 code, and 0.15% (N = 760) were linked to ME/CFS-relevant primary care codes. We therefore removed as controls those UKB participants linked to these ME/CFS-relevant codes.

We also excluded as controls those among approximately 50,000 UKB participants who were genotyped using the UK BiLEVE array, not the Axiom array. However, we will not exclude them as UKB ME/CFS cases for GWAS-3 and GWAS-4.

In addition to the genotyping quality-control that UKB performed (14), we removed variants that showed: (i) discordance between our own and UKB's genotype calls in a batch of 4,700 UKB samples, or (ii) a large departure of allele frequency in European-ancestry UKB samples from that reported in GnomAD (v2.1.1) for non-Finnish Europeans. We declared discordance when less than 95% of calls for a variant were concordant. We tested significant allele frequency differences assuming a binomial distribution of allele count, discarding any variant more than 6 standard deviations from the expected mean.

### Merged case-control array genotypes

We resolved duplicate markers and inconsistencies of chromosome, position, strand or alleles, removing variants when this was not possible. We resolved kinship and PCA-based ancestry matching and removed from further analyses any variants that were discordant in more than one pair of the 46 pairs of identical genotypes (duplicates or three pairs of monozygous twins) across datasets. We removed from the UKB cohorts those samples that were duplicated between UKB and DecodeME.

### Phasing and genotype imputation

Genotype imputation is an important step prior to any GWAS. It allows us to densify the genetic data for all individuals by inferring millions of unobserved genotypes from a whole-genome-based reference panel. The imputed genotypes boost the statistical power of a GWAS by making it more likely that we will capture the causal risk variant if there is a true association signal in a genomic region.

We imputed cases and controls together using the UKB-RAP imputation pipeline (22), with a reference panel created from the whole genome sequences of more than 200,000 UKB samples and covering over 700 million variants (23). Imputing cases and controls jointly should avoid technical batch effects that are introduced when imputing cases and controls separately, as was proposed in DecodeME's original Data Analysis Plan (11).

We only imputed samples of European PCA-based ancestry. Before we merged these samples, we filtered out variants with HWE $p < 10^{-12}$ for cases, or $< 10^{-9}$ for controls, reasoning that stronger departure from HWE might naturally occur for cases. After merging, we discarded variants that were rare (MAF < 0.01) or had a call rate < 0.98; we removed samples with genotype missingness > 3%. We then randomly subsetted control samples to match the cases' female-to-male ratio of 5:1. A total of 15,857 cases and 265,785 controls were available for imputation. We lifted over the genotypes from genome assembly build 37 (GRCh37/hg19) to build 38 (GRCh38/hg38) using the UCSC LiftOvertool, and then uploaded the data onto the UKB-RAP.

Before imputation, it was essential to perform specific quality checks relating to the imputation reference panel that we used. We ensured that the strand, alleles, position, and

REF/ALT assignments matched, and then compared the frequency differences between the genotyped data and the reference panel. We removed DNA variants if they were ambiguous (A/T or G/C, when MAF > 0.4), or had differing alleles, or were not in the reference panel. For controls, but not cases, we removed variants with an allele frequency difference between genotyped and reference genotypes of more than 10%.

Phasing is a critical step before imputation which improves its efficiency and accuracy (24). For each individual, this estimates the haplotypes – blocks of variants inherited together either from the paternal or maternal genome. We performed phasing per chromosome using the phase_common option in SHAPEIT5 (23). This implements a reference-based haplotype estimation that makes good use of the UKB reference panel. After converting the phased reference panel into the required xcf format, we undertook phasing per chromosome, on all samples together.

To make imputation easier, we divided the phased genotype data per chromosome into 26 batches. Each contained 10,832 $\pm$ 1 samples and kept the original ratio of cases to controls. We then performed imputation with IMPUTE5 (25) for each batch using the UKB reference whole genome sequencing panel. After imputation, we discarded for each batch any variant that was monomorphic, multi-allelic, or very rare (MAF < $1 \times 10^{-4}$ or minor allele count [MAC] < 20), or had low imputation quality (INFO < 0.4) (26). We then merged all batches and removed variants with MAF < $1 \times 10^{-4}$ or that had recalculated INFO < 0.4. Finally, we converted imputed genotypes into pgen format using PLINK 2.0.

### HLA imputation

We calculated HLA imputations using the HLA*IMP:02 algorithm in order to match the process done by UK Biobank (14), and as implemented by the Thermo Fisher Axiom Analysis Software (AxAS). For each batch, we removed markers within the HLA region (chromosome 6: 28,477,797-33,448,354; GRCh37/hg19) that were not best and recommended by the AxAS calling algorithm, or with low clustering metrics (as described above, in **Quality control steps and further analysis**; FLD < 4.5, Heterozygous Strength Offset < 0 or Homozygous Ratio Offset < 0). We visually inspected the remaining genotyping clusters to assess clustering quality and removed low-quality clusters (as defined above).

We exported genotypes from the 15,849 participants of European ancestry in VCF format. For each batch, we filtered out HLA markers with a call rate < 98%, using BCFtools v1.20. Then we merged the four batches into a single VCF file containing 5,412 intersected markers. We then performed further quality control steps, discarding all palindromic or multiallelic variants, and variants with HWE $p$-value < $1 \times 10^{-15}$, MAF < $6 \times 10^{-4}$ (reflecting MAC < 20) or call rate < 99% (14). We also removed all cases with data missingness > 3%.

After QC, we kept 15,841 cases and 4,912 markers for HLA imputation. For practical reasons, we split the merged data into the original batches and used the Axiom HLA analysis software for imputation. After imputation, we converted DecodeME case imputations to UKB format (one row per participant, with two imputations per locus) in *R* (27). We merged these with the UKB-provided HLA imputations, and also subsetted to include only those in the GWAS-1 cohort. This removed the outliers that we identified by manually inspecting the PCA plots (see **Ancestry and kinship**, above) (28). We used genetic dosages directly, rather than inferring genotype calls, to account for the uncertainty in these imputations (28).

As recommended in (14), we filtered UKB data to exclude imputations with posterior probabilities Q2 < 0.7 (where either an individual allele imputation, or the sum for homozygotes, was low quality). The UKB provides only the combined probabilities (Q2 scores) and not the output file of their HLA imputations. This means that information may have been lost for rare samples that are homozygous for a given HLA allele, where the Q2 scores are summed. We investigated the effects of altering our data preparation to more closely match the UKB imputations and concluded that the effects on our lead associations were minor (**Table S11**).

For DecodeME's data, the quality scores for each allele copy were available, and so we filtered them at Q < 0.7 before conversion to UKB format. We filtered samples for both DecodeME and UKB, locus-wise, if they had fewer than two imputations for that locus. We only included those samples in DecodeME that had HLA alleles reported by the UK Biobank.

### Association testing

REGENIE (29) is a machine-learning method performing whole-genome regression on both quantitative and binary phenotypes on data from a large number of individuals. The approaches implemented in REGENIE let us account for co-factors or covariates that could influence risk of ME/CFS and/or confound case-control genetic associations. These included sex and the ancestry PCs which explained most of the inter-individual variation. REGENIE provides additional fitting of a polygenic random effect that accounts for cryptic and non-cryptic relatedness. We used the Firth logistic regression implemented in REGENIE, which is robust to case-control imbalance. We performed REGENIE analysis on cases and controls of European genetic ancestry only.

We defined participants' ages as 2025 minus their year of birth. For a limited number of questionnaire fields, we removed a single participant whose recorded age of over 120 years old was an outlier. At the time of recruitment, UK Biobank volunteers were aged between 40 and 69 years old (median 56) while the DecodeME DNA participants' ages spanned from 16 to over 90 years old (median 50; mean 49). The narrower age-range among the UK Biobank controls meant that 24% of DecodeME cases could not be age-matched. This age mismatch meant that adjusting for age in the GWAS analysis would not be practicable.

We disregarded non-HLA statistical associations if they failed one or more of four QC tests:

(1) Imputation quality INFO < 0.9.

(2) Significant hits can be enriched in variants that are not in HWE (30). Nevertheless, variants that showed a large departure from HWE ($p < 1 \times 10^{-15}$) in controls needed further scrutiny. We required the HWE *p*-values of such variants to be less than $10^{-9}$ in other control populations of the same ancestry.

(3) The strength of association (*p*-value) of a variant is expected to be proportional to the genetic correlation (LD) between the variant and the SNP leading the association signal. The higher the LD correlation, the lower the *p*-value. Therefore, if an association *p*-value is inconsistent with LD then the association is likely spurious. We used LD-based QC, as implemented in DENTIST (31), to detect such likely errors in GWAS summary statistics.

(4) We inspected the genotyping cluster of any genotyped variants that were significant SNPs or that were LD-clumped with a significant lead SNP. We did this in cases and, if needed, in controls (but limited to one UKB batch only), following the criteria discussed above. For any miscalled variant that appeared to affect imputation locally, we performed a new local (chromosome-wide) imputation with the recalled or removed variant(s). Then we reran the GWAS. Finally, significant signals with a single variant (that is, without 'LD buddies') that passed all the above QC steps were flagged but not considered for post-GWAS analysis or interpretation.

*Attempted replication*

We called our primary study 'GWAS-1'. We seek to replicate its findings using UKB ME/CFS cases rather than DecodeME cases. However, in order for the replication to be independent of GWAS-1, we could not use any of the same UKB control individuals.

We therefore split the UKB controls into two unequal groups, one to be a control group for the DecodeME cases, and the other to be a control group for the UKB cases, to allow us to compare two independent GWAS. We assigned controls randomly but matched the case sex-ratio (5:1) in both the DecodeME and UKB case groups.

The resulting new DecodeME GWAS ('GWAS-2') had the same design as GWAS-1 and used all of the same DecodeME cases, but it had a lower (1:10) ratio of DecodeME cases to UKB controls (15,579 cases and 155,790 controls). This leaves enough UKB control individuals for the replication GWAS which we will report in a subsequent manuscript.

For HLA sensitivity analyses, we defined an additional cohort of those with primarily 'White British' ancestry. After performing the PCA described above, we selected as controls all remaining individuals in 'GWAS-1' that UKB had included in their definition of White British (Data Field 22006). We then used the calculated PCs to determine the geometric median (32) of only the controls, and the distance of the furthest control along each PC direction from this median. We then excluded all DecodeME case samples that were further from the median than this most extreme control. We noted from the PC plots that only the first seven PCs were likely to affect this filtering. However, to avoid normalisation across an arbitrary number of

PCs by using a distance metric, we filtered samples along each axis separately, for the first 16 PCs (20). After this filtering procedure, 14,724 cases and 230,745 controls remained. We then performed another round of PCA on this newly defined cohort, to obtain the PCs needed for UK regional-level analyses.

We used a logistic model in *R* for HLA association testing, with genetic sex and the first 20 principal components (20) as covariates. We performed sex-stratified analyses in the same way, except that we removed genetic sex as a covariate. Of the 362 alleles detected in the UKB cohort, 108 were not found among DecodeME cases, probably due to cohort size differences. Only 100 alleles had frequencies over 1%, setting our HLA-region-wide multiple testing correction threshold to $p$ < 0.05/100 (0.0005). Results are also reported at the genome-wide significance level ($p < 5 \times 10^{-8}$).

We performed a sensitivity analysis using the White British cohort for the HLA association testing, as this genomic interval has a high rate of recombination and has been shown to be associated with regional-level ancestries (33). We therefore used the same logistic model and tests as previously described, except with 72 PCs fitted instead of 20 (34).

### Replication analyses

To explore replication, we performed two analyses using fixed-effects meta-analysis as implemented in METAL (35) and PLINK 1.9 (36). The first meta-analysis, R-1, compared the genotypes of people with evidence of fatigue and post-exertional malaise, but not necessarily with an ME/CFS diagnosis, obtained from Lifelines (37) (3,440 cases and 17,080 controls) or the UKB (10,327 cases and 195,103 controls). In Lifelines, post-exertional malaise was defined when a participant answered one or more of questions 14-18, and 75-77 of the DePaul Symptom Questionnaire 2 (38), indicating at least "about half the time" with at least "moderate severity". UKB cases were defined as those who answered 'Yes' to the pain questionnaire field 120114 ('Persistent or recurrent tiredness, weariness or fatigue that has lasted for at least 6 months'), 'No' to field 120115 ('Tiredness, weariness or fatigue goes away when resting') and 'Yes' to field 120117 ('Tired after minimal physical or mental exertion'). This analysis used METAL (35).

The second meta-analysis, R-2, used cases defined by electronic health records (ICD10:G93.3 code in FinnGen, the Estonian Biobank, and the Mass General Brigham Biobank, or chronic fatigue syndrome Phecode Phe_798_1 in the Michigan Genomics Initiative and the Million Veteran Program). We defined controls as everyone not meeting the case definition. The electronic health record data in R-2 comprised that from the Estonian Biobank (39) (1,926 cases, 195,103 controls), FinnGen (40) (283 cases, 463,029 controls), the Michigan Genomics Initiative (41) (Freeze 6: 3,926 cases, 57,247 controls), the Mass General Brigham Biobank (42) (114 cases, 51,055 controls), the Million Veteran Program (43) (4,948 cases, 617,301 controls), and Genes and Health (44) (3,053 'chronic fatigue' cases, defined in part by

ICD10:G93.3 and ME/CFS SNOMED codes, and 49,755 controls). This analysis used PLINK 1.9 (36).

### Post-GWAS analyses

To visualise associations, we used LocusZoom, an online plotting platform (45). It generates both quantile-quantile (QQ) and Manhattan plots that are contextual, showing gene annotations and local linkage disequilibrium patterns. For functional mapping and to annotate genetic associations we used FUMA (https://fuma.ctglab.nl/) (46). We used the FUMA-provided data sources for gene expression QTL (eQTL) mapping. We also tested for positive relationships between gene expression in multiple tissue types and ME/CFS genetic associations using MAGMA (47), as implemented in FUMA. We also interrogated OpenTargets Genetics (48).

We used the following chromosomal intervals defined by FUMA (GRCh38): chr1:173714815-173922679, chr6:26180406-26338469, chr6:97901488-98099429, chr13:53103588-53289469, chr15:54845408-54892346, chr17:52147538-52264626 and chr20:48895255-49297643.

**Table S10. Primary care codes.** Primary care CTV3 and ReadV2 codes for diagnoses associated with ME, CFS or post-viral fatigue syndrome

| Code | Diagnostic |
|---|---|
| F03y. | Other causes of encephalitis (& [myalgic encephalomyelitis] or [encephalomyelitis NOS]) Other causes of encephalitis Encephalomyelitis NOS Myalgic encephalomyelitis |
| XE17Z | Postinfective encephalitis (& [myalgic encephalitis] or [myalgic encephalomyelitis]) Post-infectious encephalitis Post-infectious encephalitis Myalgic encephalitis Postinfective encephalitis |
| XE17b | Encephalitis/myelitis: [NOS] or [encephalomyelitis & (myalgic)] Encephalomyelitis Myalgic encephalomyelitis Encephalitis/myelitis NOS |
| .F12Z | Encephalitis, myelitis and encephalomyelitis Chronic fatigue syndrome |
| Xa01F | Chronic fatigue syndrome Myalgic encephalomyelitis ME Myalgic encephalomyelitis Myalgic encephalomyelitis syndrome Postviral fatigue syndrome PVFS - Postviral fatigue syndrome CFS - Chronic fatigue syndrome |
| .F122 | Postinfective encephalitis (& [myalgic encephalitis] or [myalgic encephalomyelitis]) Post-infectious encephalitis Myalgic encephalomyelitis Myalgic encephalitis Postinfective encephalitis |
| .F38. | Chronic fatigue syndrome Myalgic encephalomyelitis ME - Myalgic encephalomyelitis Myalgic encephalomyelitis syndrome Postviral fatigue syndrome PVFS - Postviral fatigue syndrome CFS - Chronic fatigue syndrome |
| F286. | Chronic fatigue syndrome Myalgic encephalomyelitis Myalgic encephalomyelitis ME - Myalgic encephalomyelitis Myalgic encephalomyelitis syndrome Postviral fatigue syndrome PVFS - Postviral fatigue syndrome PVFS - Postviral fatigue syndrome CFS - Chronic fatigue syndrome |
| X75s8 | Chronic fatigue syndrome Myalgic encephalomyelitis ME - Myalgic encephalomyelitis Myalgic encephalomyelitis syndrome Postviral fatigue syndrome PVFS - Postviral fatigue syndrome CFS - Chronic fatigue syndrome |
| XM06p | Chronic fatigue syndrome Myalgic encephalomyelitis ME Myalgic encephalomyelitis Myalgic encephalomyelitis syndrome Postviral fatigue syndrome PVFS - Postviral fatigue syndrome CFS - Chronic fatigue syndrome |
| F2860 | Mild chronic fatigue syndrome |
| F2861 | Moderate chronic fatigue syndrome |
| F2862 | Severe chronic fatigue syndrome |
| XaPom | Mild chronic fatigue syndrome |
| XaPon | Moderate chronic fatigue syndrome |
| XaPoo | Severe chronic fatigue syndrome |
| | **Activity management** |
| XaPeC | Activity management for chronic fatigue syndrome Activity management for myalgic encephalopathy Actvty managm for myalg enceph |
| .8Q1. | Activity management for chronic fatigue syndrome Activity management for myalgic encephalopathy Actvty managm for myalg enceph |
| 8Q1.. | Activity management for chronic fatigue syndrome Activity management for myalgic encephalopathy Actvty managm for myalg enceph |
| | **Referrals** |

| Code | Diagnostic |
|---|---|
| XaR7C | Referral to chronic fatigue syndrome specialist team Referral to myalgic encephalomyelitis specialist team |
| XaRAz | Referral for chronic fatigue syndrome activity management Referral for myalgic encephalopathy activity management |
| 8HIL. | Referral for chronic fatigue syndrome activity management Referral for myalgic encephalopathy activity management |
| 8HkW. | Referral to chronic fatigue syndrome specialist team Referral to myalgic encephalomyelitis specialist team |

**Table S12: DecodeME Participant Numbers across the Workflow**

| Category | Number |
|---|---|
| **SIGN-UPS** | |
| **Total sign-ups** | **37,767** |
| Sign-ups who were sent paper questionnaire | 759 |
| Sign-ups who completed paper questionnaire | 596 |
| Duplicate sign-up accounts deleted | 358 |
| Sign-ups who withdrew from the study | 292 |
| People who completed sign-up but declined consent | 82 |
| **SIGN-UP COMPLETERS AND CONSENTERS (‘PARTICIPANTS’)** | |
| **Total participants who completed sign-up and consented to take part in the study** | **26,901** |
| Post-Covid participants | 1,146 |
| Participants who gave permission to be sent study updates | 26,437 |
| Participants who gave permission for access to their NHS data | 23,828 |
| Participants who gave permission for other researchers to access their data | 23,055 |
| Participants who gave permission to be contacted by other studies | 25,558 |
| **DNA PARTICIPANTS (‘CASES’)** | |
| **Total participants who passed the case eligibility criteria (‘cases’)** | **21,620** |
| Post-Covid cases | 729 |
| Cases who gave permission to be sent study updates | 21,329 |
| Cases who gave permission for access to their NHS data | 19,207 |
| Cases who gave permission for other researchers to access their data | 18,674 |
| Cases who gave permission to be contacted by other studies | 20,691 |
| **SALIVA SAMPLE KITS AND EXTRACTED SAMPLES** | |
| **Total number of saliva sample kits sent** | **23,394** |
| First kits | 21,658 |
| Subsequent kits | 1,736 |
| **Returned kits (including for cases who returned a second kit)** | **18,981** |
| Cases with returned kits | 18,068 |
| **Total number of samples whose extraction was attempted** | **22,215** |
| First attempted extractions | 22,071 |
| Second attempted extractions | 3,109 |
| ≥ 60 ng/uL | 18,660 |
| 50–60 ng/uL (second extractions plus those considered for Thermo Fisher) | 663 |
| < 50 ng/uL (failed for GWAS but are stored) | 2,582 |
| Negative values, N/A entries and nulls | 310 |
| **SAMPLES SENT TO THERMO FISHER** | |
| **Samples sent for GWAS to Thermo Fisher** | **18,347** |
| Samples genotyped by Thermo Fisher | 18,266 |
| Genotyped samples that passed Thermo Fisher calling | 17,573 |
| **Genotyped samples that passed DecodeME calling** | **17,519** |
| Samples with sex inferred from DNA as ‘unknown’ | 988 |
| Samples recovered from ‘unknown sex’ | 973 |
| Samples with sex-chromosome aneuploidy or mosaicism | 11 |
| **Genotyped samples that passed DecodeME calling quality control** | **17,504** |

### REFERENCES


1. Webster-Philp O, McDonald N. The history of M.E. CFS, the evolution and the role of the CMRC (a patient's perspective). ME/CFS Alliance Think Tank; 2020.
2. Carruthers BM et al. Myalgic Encephalomyelitis/Chronic Fatigue Syndrome. Clinical Working Case Definition, Diagnostic and Treatment Protocols. Journal of Chronic Fatigue Synderome. 2003;11(1).
3. NIHR. UK Standards for Public Involvement [Internet]. [cited 2024 Dec 5]. Available from: https://sites.google.com/nihr.ac.uk/pi-standards/home
4. Devereux-Cooke A, Leary S, McGrath SJ, Northwood E, Redshaw A, Shepherd C, et al. DecodeME: community recruitment for a large genetics study of myalgic encephalomyelitis / chronic fatigue syndrome. BMC Neurol. 2022 Jul 19;22(1):269.
5. Devereux-Cooke A. Representing public voices in a research study [Internet]. [cited 2025 Jul 24]. Available from: https://www.ukri.org/manage-your-award/good-research-resource-hub/research-co-production/representing-public-voices-in-a-research-study/
6. Gov.uk. My full reality: the interim delivery plan on ME/CFS [Internet]. [cited 2024 Dec 5]. Available from: https://www.gov.uk/government/consultations/improving-the-experiences-of-people-with-mecfs-interim-delivery-plan/my-full-reality-the-interim-delivery-plan-on-mecfs
7. Pendergrast T, Brown A, Sunnquist M, Jantke R, Newton JL, Strand EB, et al. Housebound versus nonhousebound patients with myalgic encephalomyelitis and chronic fatigue syndrome. Chronic Illn. 2016 Dec;12(4):292–307.
8. Committee on the Diagnostic Criteria for Myalgic Encephalomyelitis/ Chronic Fatigue Syndrome. Beyond Myalgic Encephalomyelitis/Chronic Fatigue Syndrome: Redefning an Illness. Washington, DC: The National Academies Press. 2015;22015.
9. 25% M.E. Group. 25% M.E Group [Internet]. [cited 2024 Dec 5]. Available from: https://test1.25megroup.org/
10. The Marmot Review. Fair Society Healthy Lives [Internet]. 2010 [cited 2025 Apr 22]. Available from: https://www.instituteofhealthequity.org/resources-reports/fair-society-healthy-lives-the-marmot-review/fair-society-healthy-lives-full-report-pdf.pdf
11. DecodeME Genetics Delivery Team. Data Analysis Plan. 2023 [cited 2025 Apr 22]; Available from: https://www.decodeme.org.uk/app/uploads/2023/03/DecodeME-GWAS-Analysis-Plan-v1.pdf
12. DecodeME Genetics Delivery Team. Data Analysis Plan [Internet]. 2024 [cited 2025 Apr 22]. Available from: https://www.decodeme.org.uk/app/uploads/2024/03/2024-03-15_DecodeME_Data_Analysis_Plan_v2_final.pdf
13. Thermofisher. Axiom™ Genotyping Solution Data Analysis USER GUIDE [Internet]. MAN0018363. 2020 [cited 2024 Nov 1]. Available from: https://assets.thermofisher.com/TFS-Assets/LSG/manuals/axiom_genotyping_solution_analysis_guide.pdf

14. Bycroft C, Freeman C, Petkova D, Band G, Elliott LT, Sharp K, et al. The UK Biobank resource with deep phenotyping and genomic data. Nature. 2018 Oct;562(7726):203–9.

15. 1000 Genomes project Consortium. IGSR: The International Genome Sample Resource. [Internet]. 2015 [cited 2025 Jul 24]. Available from: https://www.internationalgenome.org/

16. Forgetta V, Li R, Darmond-Zwaig C, Belisle A, Balion C, Roshandel D, et al. Cohort profile: Genomic data for 26 622 individuals from the Canadian Longitudinal Study on Aging (CLSA). BMJ Open. 2022 Mar 10;12(3).

17. Prive F, Aschard H, Ziyatdinov A, Blum MGB. Efficient analysis of large-scale genome-wide data with two R packages: Bigstatsr and bigsnpr. Bioinformatics. 2018 Aug 15;34(16):2781–7.

18. Manichaikul A, Mychaleckyj JC, Rich SS, Daly K, Sale M, Chen WM. Robust relationship inference in genome-wide association studies. Bioinformatics. 2010 Nov;26(22):2867–73.

19. 2024 [Internet]. [cited 2025 Jul 24]. Axiom™ Analysis Suite v5.4 User Guide. Available from: https://assets.thermofisher.com/TFS-Assets/GSD/brochures/axiom-analysis-suite-user-guide.pdf

20. Privé F, Luu K, Blum MGB, McGrath JJ, Vilhjálmsson BJ. Efficient toolkit implementing best practices for principal component analysis of population genetic data. Bioinformatics. 2020 Aug 15;36(16):4449–57.

21. Allen NE, Lacey B, Lawlor DA, Pell JP, Gallacher J, Smeeth L, et al. Prospective study design and data analysis in UK Biobank. Sci Transl Med. 2024 Jan 10;16(729):eadf4428.

22. Rubinacci S, Delaneau O. 2024. [cited 2025 Jul 24]. UK Biobank imputation pipelines. Available from: https://srubinacci.gitbook.io/uk-biobank-imputation-pipelines

23. Hofmeister RJ, Ribeiro DM, Rubinacci S, Delaneau O. Accurate rare variant phasing of whole-genome and whole-exome sequencing data in the UK Biobank. Nat Genet. 2023 Jul 1;55(7):1243–9.

24. Howie B, Fuchsberger C, Stephens M, Marchini J, Abecasis GR. Fast and accurate genotype imputation in genome-wide association studies through pre-phasing. Nat Genet. 2012 Aug;44(8):955–9.

25. Rubinacci S, Delaneau O, Marchini J. Genotype imputation using the Positional Burrows Wheeler Transform. PLoS Genet. 2020 Nov 16;16(11).

26. Howie B, Marchini J, Stephens M. Genotype imputation with thousands of genomes. G3: Genes, Genomes, Genetics. 2011 Nov;1(6):457–70.

27. R Core Team. R: A Language and Environment for Statistical Computing [Internet]. 2024 [cited 2025 Apr 22]. Available from: https://www.r-project.org/

28. Sakaue S, Gurajala S, Curtis M, Luo Y, Choi W, Ishigaki K, et al. Tutorial: a statistical genetics guide to identifying HLA alleles driving complex disease. Vol. 18, Nature Protocols. Springer Nature; 2023. p. 2625–41.

29. Mbatchou J, Barnard L, Backman J, Marcketta A, Kosmicki JA, Ziyatdinov A, et al. Computationally efficient whole-genome regression for quantitative and binary traits. Nat Genet. 2021 Jul 1;53(7):1097–103.

30. Truong VQ, Woerner JA, Cherlin TA, Bradford Y, Lucas AM, Okeh CC, et al. Quality Control Procedures for Genome-Wide Association Studies. Curr Protoc. 2022 Nov;2(11):e603.

31. Chen D, Tashman K, Palmer DS, Neale B, Roeder K, Bloemendal A, et al. A data harmonization pipeline to leverage external controls and boost power in GWAS. Hum Mol Genet. 2022 Feb 3;31(3):481–9.

32. Privé F, Aschard H, Carmi S, Folkersen L, Hoggart C, O'Reilly PF, et al. Portability of 245 polygenic scores when derived from the UK Biobank and applied to 9 ancestry groups from the same cohort. Am J Hum Genet. 2022 Jan 6;109(1):12–23.

33. Winney B, Boumertit A, Day T, Davison D, Echeta C, Evseeva I, et al. People of the British Isles: preliminary analysis of genotypes and surnames in a UK-control population. Eur J Hum Genet. 2012 Feb;20(2):203–10.

34. Abdellaoui A, Hugh-Jones D, Yengo L, Kemper KE, Nivard MG, Veul L, et al. Genetic correlates of social stratification in Great Britain. Nat Hum Behav. 2019 Dec 1;3(12):1332–42.

35. Willer CJ, Li Y, Abecasis GR. METAL: fast and efficient meta-analysis of genomewide association scans. Bioinformatics. 2010 Sep 1;26(17):2190–1.

36. Chang Christopher C, Chow Carson C, Tellier Laurent C, Vattikuti Shashaank, Purcell Shaun M, Lee James J. Second-generation PLINK: rising to the challenge of larger and richer datasets. Gigascience. 2015;4(7).

37. ME/CFS Lines: A multidisciplinary consortium & biobank to unravel the causes of ME/CFS in Lifelines [Internet]. 2025 [cited 2025 Jul 24]. Available from: https://projecten.zonmw.nl/en/project/mecfs-lines-multidisciplinary-consortium-biobank-unravel-causes-mecfs-lifelines

38. Bedree H, Sunnquist M, Jason LA. The DePaul Symptom Questionnaire-2: A Validation Study. Fatigue. 2019;7(3):166–79.

39. Leitsalu L, Haller T, Esko T, Tammesoo ML, Alavere H, Snieder H, et al. Cohort Profile: Estonian Biobank of the Estonian Genome Center, University of Tartu. Int J Epidemiol. 2015 Aug;44(4):1137–47.

40. Kurki MI, Karjalainen J, Palta P, Sipilä TP, Kristiansson K, Donner KM, et al. FinnGen provides genetic insights from a well-phenotyped isolated population. Nature. 2023 Jan;613(7944):508–18.

41. Zawistowski M, Fritsche LG, Pandit A, Vanderwerff B, Patil S, Schmidt EM, et al. The Michigan Genomics Initiative: A biobank linking genotypes and electronic clinical records in Michigan Medicine patients. Cell genomics. 2023 Feb 8;3(2):100257.

42. Karlson EW, Boutin NT, Hoffnagle AG, Allen NL. Building the Partners HealthCare Biobank at Partners Personalized Medicine: Informed Consent, Return of Research Results, Recruitment Lessons and Operational Considerations. J Pers Med. 2016 Jan 14;6(1).

43. Verma A, Huffman JE, Rodriguez A, Conery M, Liu M, Ho YL, et al. Diversity and scale: Genetic architecture of 2068 traits in the VA Million Veteran Program. Science. 2024 Jul 19;385(6706):eadj1182.

44. Finer S, Martin HC, Khan A, Hunt KA, MacLaughlin B, Ahmed Z, et al. Cohort Profile: East London Genes & Health (ELGH), a community-based population genomics and health study in British Bangladeshi and British Pakistani people. Int J Epidemiol. 2020 Feb 1;49(1):20–21i.

45. Boughton AP, Welch RP, Flickinger M, Vandehaar P, Taliun D, Abecasis GR, et al. LocusZoom.js: interactive and embeddable visualization of genetic association study results. Bioinformatics. 2021 Sep 15;37(18):3017–8.

46. Watanabe K, Taskesen E, Van Bochoven A, Posthuma D. Functional mapping and annotation of genetic associations with FUMA. Nat Commun. 2017 Dec 1;8(1).

47. de Leeuw CA, Mooij JM, Heskes T, Posthuma D. MAGMA: Generalized Gene-Set Analysis of GWAS Data. PLoS Comput Biol. 2015 Apr 1;11(4).

48. Mountjoy E, Schmidt EM, Carmona M, Schwartzentruber J, Peat G, Miranda A, et al. An open approach to systematically prioritize causal variants and genes at all published human GWAS trait-associated loci. Nat Genet [Internet]. 2021;53(11):1527–33. Available from: https://www.nature.com/articles/s41588-021-00945-5
